# Longitudinal peripheral blood multi-omic profiling in seropositive individuals identifies immune endotypes and predictive models for future rheumatoid arthritis conversion

**DOI:** 10.64898/2026.02.12.26346058

**Authors:** Jun Inamo, Aleksandra Bylinska, Miles Smith, Lauren Vanderlinden, Christian Wright, Tayte Stephens, Marie L. Feser, Christopher C. Striebich, James R. O’Dell, Jeffrey A. Sparks, John M. Davis, Jonathan Graf, Maureen A. McMahon, E. Blair Solow, Lindsy J. Forbess, Athan N. Tiliakos, David A. Fox, Maria I. Danila, Diane Lewis Horowitz, Jonathan Kay, Judith A. James, V. Michael Holers, Kevin D. Deane, Joel M. Guthridge, Fan Zhang

**Affiliations:** Division of Rheumatology, University of Colorado School of Medicine, Aurora, CO, United States of America; Department of Biomedical Informatics, Center for Health Artificial Intelligence, University of Colorado School of Medicine, Aurora, CO, United States of America; Arthritis & Clinical Immunology Research Program, Oklahoma Medical Research Foundation, Oklahoma City, OK, United States of America; University of Nebraska Medical Center, Omaha, NE, USA; Mass General Brigham, Brigham and Women’s Hospital and Harvard Medical School, Boston, MA, USA; Division of Rheumatology, Mayo Clinic, Rochester, MN, USA; University of California San Francisco, San Francisco, CA, USA; University of California Los Angeles, Los Angeles, CA, USA; UT Southwestern Medical Center, Dallas, TX, USA; Cedars-Sinai Medical Center, Los Angeles, CA, USA; Emory University, Atlanta, GA, USA; University of Michigan, Ann Arbor, MI, USA; University of Alabama at Birmingham, Birmingham, AL, USA; Northwell Health, New Hyde Park, NY, USA; UMass Chan Medical School, Worcester, MA, USA

## Abstract

Individuals who have serum elevations of anti-cyclic citrullinated protein (anti-CCP) antibodies are at risk for developing rheumatoid arthritis (RA), yet immunologic factors that lead to a transition from pre- to clinical RA remain unclear. Here, we used materials from anti-CCP antibody-positive individuals enrolled in a clinical trial that evaluated the efficacy of hydroxychloroquine to prevent clinical RA, and performed multi-modal single-cell profiling (transcriptome, surface proteins, T/B-cell receptor sequencing, and chromatin accessibility) on samples obtained at baseline and at RA onset in those who developed clinical RA (Converters) or follow-up point in matched Nonconverters. At both baseline and follow-up, Converters had expansions of peripheral helper T (Tph) cells and CD8^+^ T cells expressing *GZMK* and *GZMB*, along with elevated potentially autoreactive T-cell receptors in CD4^+^ T cells compared to Nonconverters. Induction of age-associated B cell signatures was observed in B cells of Converters prior to RA onset. Epigenetic profiling further identified chromatin accessibility changes in Converters over time, particularly within myeloid and NK cells. Lastly, predictive modeling using baseline immune features, including Tph cells, *GZMK*^+^*XCL1*^+^ CD8^+^, and *GZMB*^+^*CD57*^+^ CD8^+^ T cells, together with clinical features such as anti-CCP3 levels, RF-positivity, and HLA shared epitope status, stratified RA risk and predicted time to onset. These findings define immune endotypes in pre-RA that could serve as targets for future preventive interventions and be used to stratify the risk of developing clinical RA in anti-CCP antibody-positive individuals.

## Introduction

Rheumatoid arthritis (RA) is a chronic autoimmune disease that develops through a stage that can be characterized by the presence of autoantibodies such as anti-cyclic citrullinated peptide (anti-CCP) antibodies and rheumatoid factor (RF), often years before the onset of clinically apparent arthritis (clinical RA)^1^. The stage prior to clinical RA, known as pre-RA, offers a unique opportunity to apply preventive interventions^2^. However, not all individuals who are anti-CCP-positive progress to clinical RA, with multiple studies demonstrating positive predictive values of anti-CCP for future clinical RA of ∼20-60%^3–5^. This heterogeneity underscores the need to elucidate the key immunologic factors that drives a transition from anti-CCP positive ‘at risk’ state to clinical RA in order to identify potential biologic targets for preventive intervention as well as to improve prediction/risk stratification for future clinical RA^6–14^.

In a previous study, we used mass cytometry and single-cell multimodal cellular indexing of transcriptomes and epitopes by sequencing (CITE-Seq) to characterize the peripheral immune landscape in a cross-sectional study of individuals at risk for clinical RA, and we identified expansions of T peripheral helper (Tph) cells, cytotoxic CD8^+^ T cells, and proinflammatory monocyte subsets^15^. An additional cross-sectional study has identified that citrullinated cartilage intermediate layer protein 1–reactive T cells^16^, HLA-DR⁺ Tph cells, and CXCR5⁻CD11c⁻CD38⁺ naive B cells^17^ are expanded in anti-CCP-positive individuals at risk for clinical RA. Furthermore, other groups have found changes in multiple immune compartments, including naive CD4⁺ T cells^18,19^, CD8⁺ T cells^20^, NK cell subsets^21^, T follicular helper (Tfh) cells^22^, and activated CD4⁺CD69^+^ T cells^23^ and CD8⁺CD69⁺ T cells^24^ that are present prior to the onset of clinical RA, and in some cases improve risk stratification for future RA. More recently, our collaborative multi-omic study longitudinally profiling anti-CCP positive at risk individuals indicated that progression to clinical RA is associated with systemic inflammation, signatures of activation in naive T and B cells, and an expansion of memory CD4⁺ T cells with signatures of B cell help^25^.

While these studies provided important insights into the temporal dynamics of the immune system in the development of RA, our overall understanding of immune alterations that precede clinical RA remains incomplete. As such, we have conducted a longitudinal study using samples from the NIH-sponsored StopRA trial^26^ (ClinicalTrials.gov NCT02603146^27^). In brief, StopRA was a phase 2, randomized, double-masked, placebo-controlled trial evaluating the efficacy of 12 months of hydroxychloroquine (HCQ) to prevent clinical RA over 36 months of follow-up. The trial population was composed of individuals who were anti-CCP3 positive at a level of ≥2 times the upper limit of normal. Although the trial demonstrated that HCQ did not prevent the development of clinical RA, we are now able to use clinical data and biospecimens collected throughout the study to evaluate baseline and longitudinal factors related to Conversion to clinical RA.

Herein, we evaluated longitudinal materials from Converters and Nonconverters from the StopRA clinical trial. To deepen our understanding from recent insights, our study employed an integrated single-cell multi-omic approach combining CITE-seq paired with T cell receptor (TCR) sequencing and B cell receptor (BCR) sequencing, mass cytometry, and simultaneous single-cell open chromatin accessibility profiling and surface protein expression (single-cell assay for transposase accessible chromatin [scATAC] with select antigen profiling by sequencing, scASAP-seq^28^). Major new findings include identification of candidate cell types and pathways involved in the transition to clinical RA, resulting in a model to predict the onset of clinical RA in anti-CCP-positive individuals.

## Results

### Study subjects

We evaluated a subset of StopRA trial participants (**Fig.1a, Supplementary Figure 1a, Supplementary Table 1**). For these analyses, we defined two primary outcome groups; Converters (n=47) who developed ‘clinical RA’ that was defined in the parent clinical trial by the fulfillment of the 2010 ACR/EULAR classification criteria for RA or the presence of ≥1 joint with swelling consistent with inflammatory arthritis (IA) and the presence of ≥1 erosion on x-rays of the hands, wrists, and feet (n=45) and an additional 2 Converters who were diagnosed with RA during the course of study at an evaluation by a clinician outside of the study, and Nonconverters who did not develop RA-like synovitis in any joint during the trial^26^. Samples from these subjects were collected at a baseline study visit for all (termed time point ‘V0’), and a study visit at the time of onset of clinical RA in Converters and at a follow-up time point in Nonconverters (‘V1’). A subset of Converters had an additional study time point evaluated prior to onset of clinical RA; this extra time point is termed ‘V0.5’.

### Longitudinal multi-modal immune profiling of anti-CCP positive individuals

We profiled 96 individuals (47 Converters, 49 Nonconverters) by mass cytometry on peripheral blood mononuclear cells (PBMCs) using the T cell Expansion and Maxpar Direct Immune Profiling Assay (MDIPA) panel (**Methods**, **Fig.1a**). For mass cytometry, we evaluated time points V0 and V1 for Converters and Nonconverters; in addition, for a subset of Converters we also evaluated time point V0.5.

For 34 individuals (24 Converters, 10 Nonconverters) who were part of the cohort for whom mass cytometry data was obtained, we also performed 137-plex CITE-seq with paired single-cell TCR and BCR sequencing, and scASAP-seq on PBMCs using time points V0 and V1 (**Supplementary Figure 1g-h**). Notably, these samples were all selected from trial participants who received placebo in order to reduce bias from drug effect..

The list of measured surface proteins across modalities are available in **Supplementary Table 2.** After quality control steps by datasets (**Methods, Supplementary Figure 1b-f**), we consistently identified four major immune compartments, including T cells, B cells, NK cells, and myeloid cells, across CITE-seq, scASAP-seq, and mass cytometry (**Fig. 1c-g**). For subsequent analyses, we included only samples corresponding to V0 and V1 and excluded V0.5 which only had mass cytometry data to ensure comparability of samples from equivalent study time points across platforms.

**Fig. 1:**
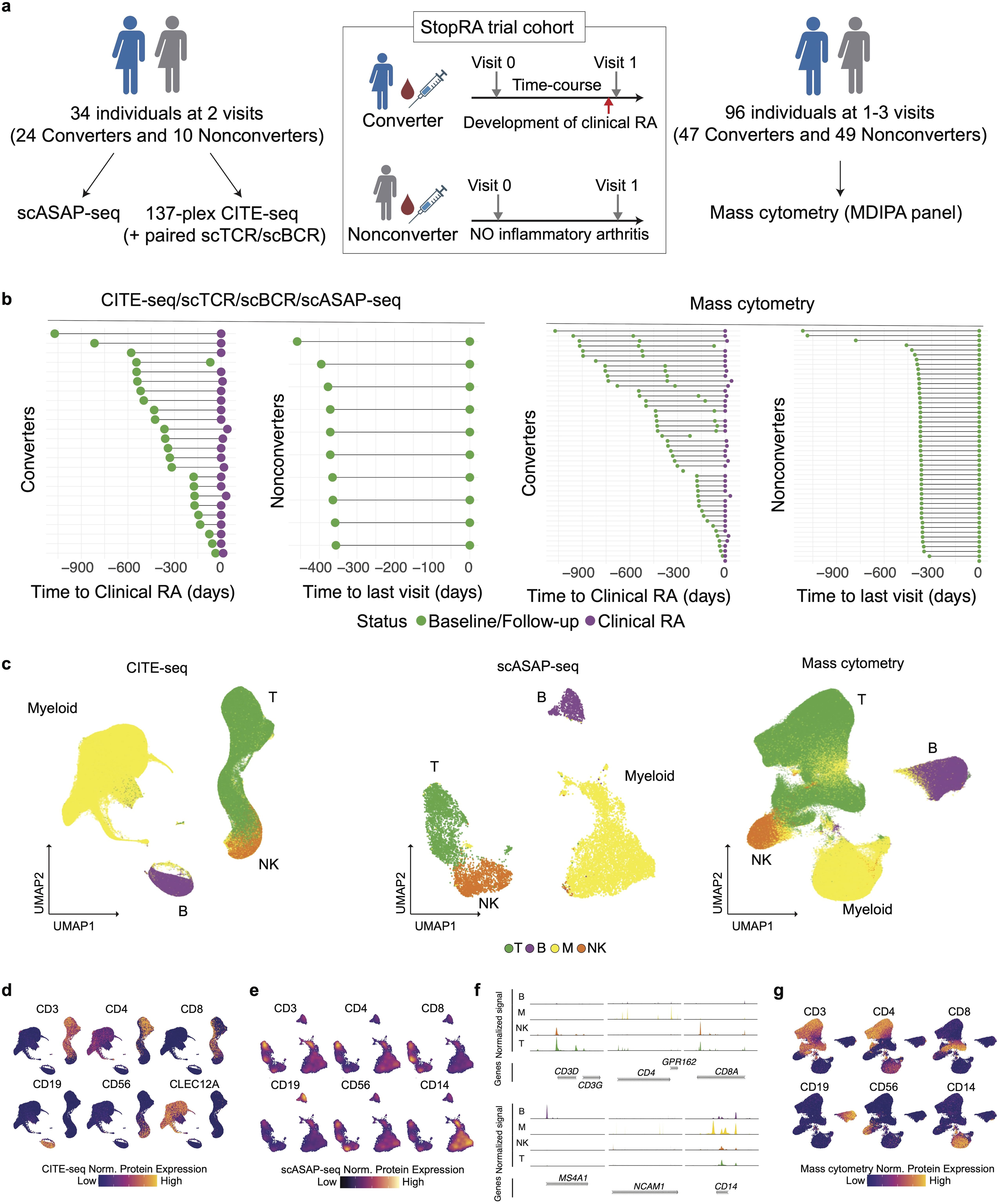
Study design for longitudinal single-cell omic data collection from RA-Converters and Nonconverters. a, Study cohort consists of a subset of individuals enrolled in the StopRA trial, with anti-CCP-positive individuals followed over multiple visits to track progression to RA. **b,** Time points of sample collection per individual. The time to clinical RA diagnosis (which is designated as day 0) is shown for Converters, while the last available visit is indicated for Nonconverters. Data are presented separately for individuals analyzed by CITE-seq/scTCR-seq/scBCR-seq/scASAP-seq (left) and those analyzed by mass cytometry (right). Colored points represent visit status: baseline/follow-up (green) or post-RA (purple). **c,** Broad immune cell-types on the UMAP space across CITE-seq, scASAP-seq, and mass cytometry datasets. **d-e,g**, Normalized protein expressions on the UMAP space in CITE-seq (**d**), scASAP-seq (**e**), and mass cytometry dataset (**g**). **f**, Fragment coverage of cell-type specific markers in scASAP-seq data.

### T cell subcluster analysis reveals early expansion of Tph and *GZMK*- and *GZMB*-expressing CD8^+^ T cell populations in individuals who develop IA

To characterize T cell dynamics preceding clinical RA, we performed an integrative analysis combining mass cytometry data from all participants with CITE-seq profiling in a representative subset (**Methods**). This approach leveraged the larger cohort coverage of mass cytometry data to define immune changes, while CITE-seq data provided deeper multi-omic annotation and transcriptional validation of key immune subsets. Together, this harmonized approach enabled robust identification of immune subsets associated with RA conversion. This joint analysis identified key CD4^+^ and CD8^+^ T cell subclusters, including peripheral helper T (Tph) cells, *CXCR5*^+^ CD4^+^ (Tfh) cells, regulatory T cells (Tregs), and cytotoxic subsets such as *GZMK*^+^ and *GZMB*^+^ CD8+ T cells (**Fig. 2a**). For details on platform-specific analyses and the integration methodology, please refer to the **Supplementary Text** and **Methods**.

**Fig. 2:**
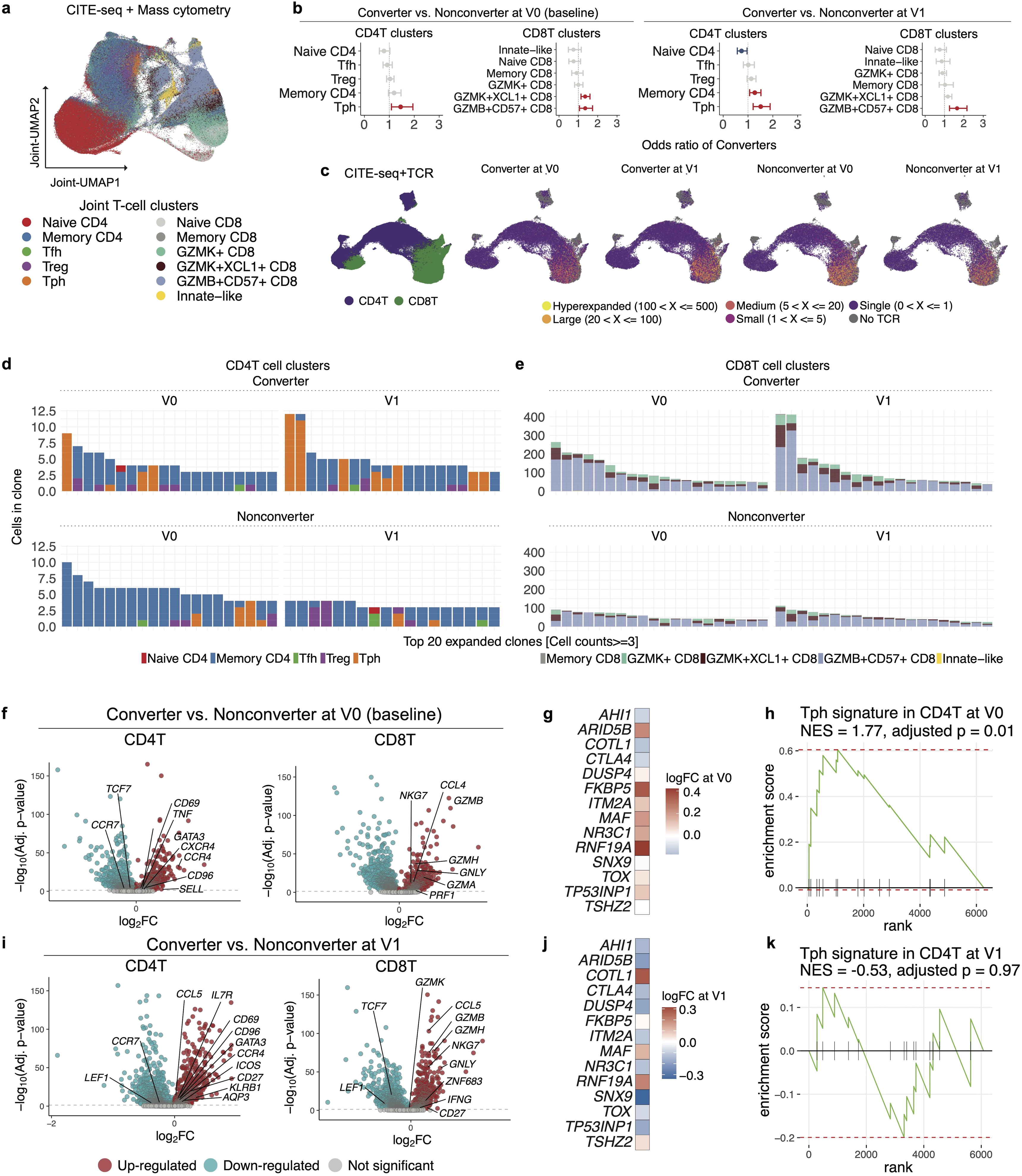
T cell subcluster analysis by integrating single-cell datasets from CITE-seq and mass cytometry identified enrichment of Tph-signature and expansion of Tph cells and *GZMK*^+^/*GZMB*^+^ CD8^+^ T cells in Converters before onset of clinical arthritis. a, T cell subclusters projected into a shared embedding space using overlapping 33 proteins measured by CITE-seq and mass cytometry data. **b,** GLMM analysis using joint T cells comparing RA Converters and Nonconverters at each visit point, at baseline (V0, left) and post-onset/follow-up (V1). The model accounted for age, sex, batch, individuals, and modalities (CITE-seq or mass cytometry) as covariates. **c**, UMAP projection of T cells in CITE-seq data colored by CD4^+^ T (purple) and CD8^+^ T cells (green), and clonal expansion of TCR repertoires across converter status and visit points. **d-e**, The composition of the top 20 expanded T cell clones is shown for CD4^+^ T cells and CD8^+^ T cells at V0 and V1. **f**, Differentially expressed genes in CD4^+^ and CD8^+^ T cells between Converters and Nonconverters at baseline (V0). T-cell marker genes are annotated. **g**, Heatmap showing expression of representative peripheral T helper (Tph) cell-signature genes comparing RA Converters and Nonconverters at baseline. **h**, Normalized enrichment score (NES) by Gene Set Enrichment Analysis (GSEA) showing the enrichment of the Tph-signature in Converter at baseline. **i**, Differentially expressed genes in CD4^+^ and CD8^+^ T cells between Converters and Nonconverters at the follow-up time point (V1). T-cell marker genes are annotated. **j**, Heatmap showing expression of representative Tph-signature genes comparing RA Converters and Nonconverters at V1. **k**, GSEA for Tph-signature at V1.

Using generalized linear mixed models (GLMMs), we tested each subcluster for association with eventual clinical RA development, while adjusting for technical batch, inter-individual differences, and technology effects (CITE-seq or mass cytometry) (**Methods**); GLMM analyses confirmed enrichment of Tph, *GZMK*^+^*XCL1*^+^ and *GZMB*^+^*CD57*^+^ CD8^+^ T cell subsets in Converters at both baseline (OR [95%CI] = 1.45 [1.08-1.95] for Tph cells; OR [95%CI] = 1.35 [1.14-1.60] for *GZMK*^+^*XCL1*^+^ CD8^+^ T cell; OR [95%CI] = 1.35 [1.06-1.73] for *GZMB*^+^*CD57*^+^ CD8^+^ T cell) and follow-up (OR [95%CI] = 1.51 [1.21-1.89] for Tph cells; OR [95%CI] = 1.27 [1.07-1.52] for memory CD4^+^ T cells; OR [95%CI] = 1.65 [1.26-2.16] for *GZMB*^+^*CD57*^+^ CD8^+^ T cell) (**Fig. 2b**) as well as comparison using frequencies (**Supplementary Figure** □**3a**).

To identify specific T cell subsets that have expanded over time, we further analyzed T cell clonal expansion in the subset of Converters and Nonconverters that had TCR sequencing data (**Supplemental Table 1b**). UMAP visualization highlighted clonally expanded T cells, in particular CD8^+^ compartments (**Fig. 2c**). The top 20 expanded CD4^+^ clones were enriched for Tph subsets in Converters at both baseline and after RA onset (**Fig. 2d**). Expanded CD8^+^ clones were predominantly composed of *GZMK*^+^ and *GZMB*^+^ subsets in Converters (**Fig. 2e**).

To assess qualitative transcriptional differences beyond cluster-level frequency shifts, we performed differential gene expression analysis on both CD4^+^ and CD8^+^ T cells using CITE-seq data. At baseline, CD4^+^ T cells from Converters showed upregulation of several differentially expressed genes (DEGs) associated with effector function and migration, including *GATA3*, *CXCR4*, *CCR4*, and *SELL* (**Fig. 2f**, left). Given our finding in the previous work of increased Tph cell frequencies in individuals at risk for RA onset^15^, we performed gene set enrichment analysis (GSEA) using a curated Tph gene signature^29^ to evaluate whether a transcriptional Tph program was present at baseline. This analysis revealed significant enrichment of the Tph signature in CD4^+^ T cells from Converters at baseline (normalized enrichment score [NES] = 1.77, adjusted *p*-value = 0.01; **Fig. 2g-h**), indicating that Tph-related transcriptional changes precede RA onset. At the follow-up time point, while CD4^+^ T cells in Converters showed elevated expression of *ICOS*, which is associated with Tph cells but also broadly reflects co-stimulatory activation states, GSEA of the Tph signature no longer reached statistical significance (NES = -0.53, adjusted *p*-value = 0.97; **Fig. 2i-k**). Instead, we observed an enrichment of the *CD96^high^* Th22-like CD4^+^ T cell signature at follow-up in Converters, suggesting the post-onset emergence of this distinct inflammatory subset (NES = 1.53, adjusted *p*-value = 0.05; **Supplementary Figure** □**3b-d**). In CD8^+^ T cell clusters, DEGs at baseline included increased expression of cytotoxicity-associated genes such as *GZMB*, *GNLY*, *NKG7*, and *GZMH* in Converters (**Fig. 2f**, right), which remained upregulated at the follow-up time point (**Fig. 2i**, right).

While these cross-sectional analyses revealed a significant difference between Converters and Nonconverters, comparisons within the Converters (V0 vs. V1) showed no significant changes in T cell subsets including Tph and activated CD8 T cells (**Supplementary Figure**□**3e-f**). This could indicate that the immunologic divergence we observed at baseline between Converters and Nonconverters was already present prior to the initial samples that we have available from the trial; alternatively, inter-individual variability might have masked longitudinal trends.

Together, these results demonstrate that the phenotypic and clonal expansion of Tph and *GZMK/GZMB*-expressing CD8^+^ T cells is present in Converters prior to incident clinical RA, and distinguishes them from Nonconverters.

### TCR repertoire analysis reveals clonal contraction in *GZMK^+^* CD8^+^ T cells and preserved diversity in CD4^+^ T cells of Converters

To characterize the TCR repertoire dynamics in Converters, we analyzed clonal diversity, measured by Hill diversity indices (*D*)^30^, across T cell subclusters. Overall, TCR diversity showed substantial variation across T cell subsets (**Supplementary Figure** □**5a**), with memory CD4^+^ and naive CD4^+^ T cells displaying the highest diversity indices. Regardless of parameter *q*, which is the parameter for down-weighting rare clones, CD4^+^ T cells retained high diversity while clonal contraction was apparent among CD8^+^ T cell subsets (**Supplementary Figure** □**5b-c**). Stratification by disease outcome and time points revealed a notable reduction in clonal diversity within *GZMK*^+^ and *GZMK*^+^*XCL1*^+^ CD8^+^ T cells in Converters after RA onset compared to baseline (**Fig. 3a**). In contrast, CD4^+^ T cell subsets maintained higher clonal diversity in Converters at both baseline and follow-up. These findings suggest distinct immunological dynamics, where expanded CD8^+^ T cells in Converters undergo clonal narrowing during RA development, while CD4^+^ T cells retain broader repertoire diversity, potentially maintaining helper functions for a wide range of antigens throughout the transition to clinical disease.

**Fig. 3:**
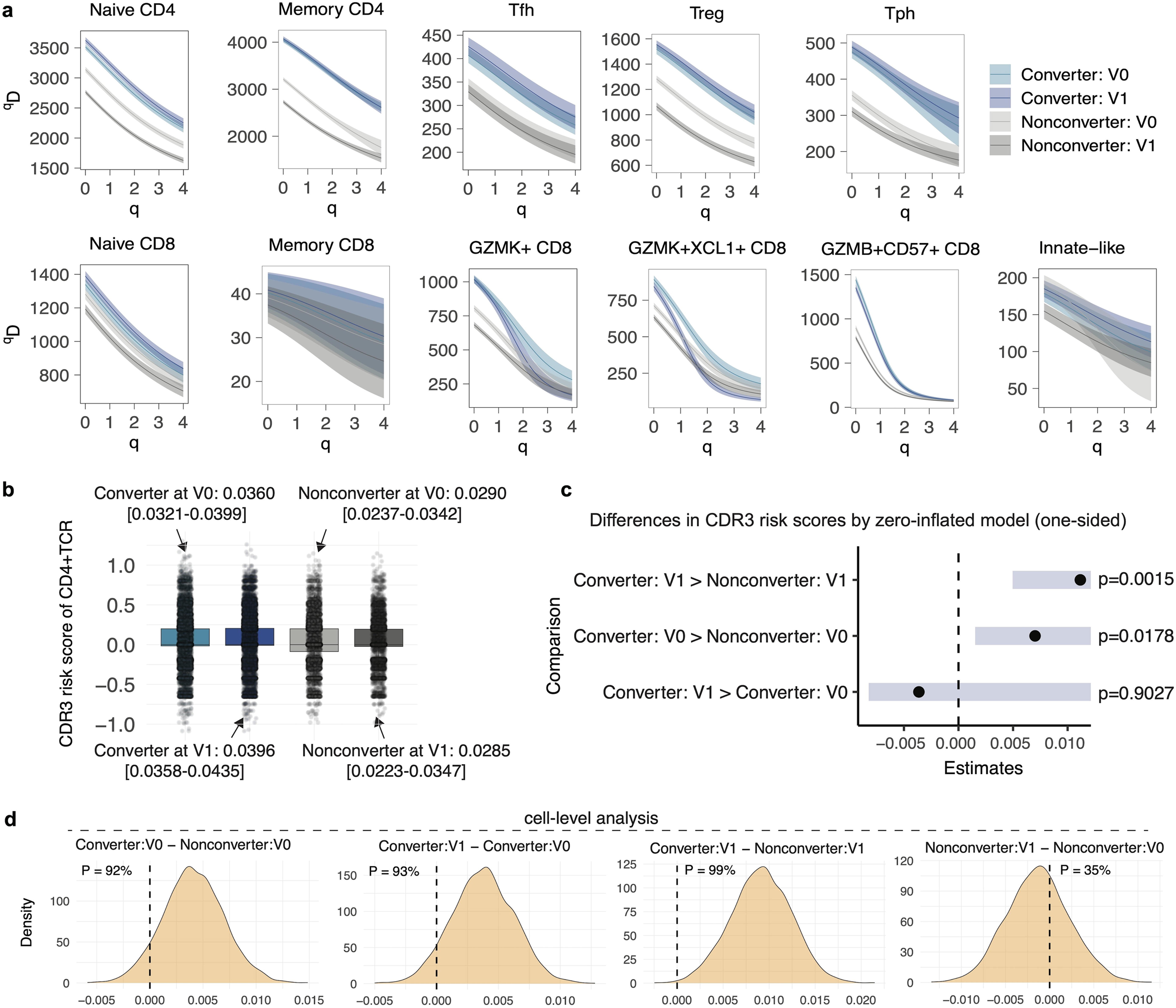
TCR analysis revealed diverse repertoire across CD4^+^ T cells and convergence in expanded CD8^+^ T cell subsets. **a**, TCR repertoire diversity by converter status and visit points in each T cell subcluster. **b**, Distribution of RA-CDR3 risk scores of CD4^+^ TCRs. Each dot represents a single-TCR. Within each boxplot, the horizontal lines reflect the median, the top and bottom of each box reflect the interquartile range (IQR), and the whiskers reflect the maximum and minimum values within each grouping no further than 1.5 × IQR from the hinge. **c**, Estimated differences in CDR3 risk scores between pairs of groups based on one-sided contrast tests from a zero-inflated Gaussian mixed model. The error bars (blue shade) represent the standard error of the estimated differences. A vertical dashed line at zero indicates the null hypothesis of no difference. Contrasts were set up under the hypothesis that the first group in each comparison has a higher mean than the second (e.g., Converter: V1 > Converter: V0). **d**, Cell-level posterior comparisons of RA-CDR3 scores between Converters and Nonconverters at baseline and follow-up. Each panel shows the posterior distribution of the difference between group means based on Bayesian modeling of single-cell RA-CDR3 scores accounting for variation by CDR3 length. Vertical dashed lines denote zero, and shaded densities represent the estimated distribution of group differences. The posterior probability P shown in each panel indicates the proportion of MCMC samples where the left group has higher scores than the right group.

### CD4^+^ TCR risk score for RA increased over time in Converters

The strongest known genetic risk for RA is the ‘shared epitope’ which comprises a number of alleles located within the human leukocyte antigen (HLA) class II region^31^. CD4^+^ T cells recognize antigen–HLA class II complexes via the complementarity-determining region 3 (CDR3) of their TCRs, leading to the selection of autoreactive T cell populations^32,33^. Therefore, to assess the emergence of autoreactive potential of CD4^+^ T cells prior to the onset of clinical RA, we applied previously established RA-associated CDR3 risk scores (RA-CDR3 scores)^33^ to our single-cell TCR repertoire data (**Fig. 3b**). At the single-cell level, we modeled the data using a zero-inflated Gaussian mixed model to account for the high frequency of zero scores, which result from the scoring method assigning non-zero coefficients only to CDR3 amino acid (AA) residues that are statistically associated with RA (**Fig. 3c**). This model confirmed that RA-CDR3 scores in Converters were significantly higher than in Nonconverters at baseline (one-sided *p*-value = 0.0178) and follow-up (one-sided *p*-value = 0.0015), while no significant increase was observed between baseline and follow-up in Converters (one-sided *p*-value = 0.9027).

We next asked whether RA-CDR3 risk signals were detectable at the individual level by stratifying CD4^+^ T cells into major phenotypic subsets. Focusing on the most abundant CD4^+^ T cell phenotypes, including naïve and memory cells, we analyzed sample-level mean RA-CDR3 scores across different CDR3 AA lengths (**Supplementary Figure 5d**). Although some variability was observed depending on CDR3 AA length, possibly due to sparse nature of single-cell data, Converters predominantly exhibited higher scores in both naïve and memory CD4^+^ T cells for selected lengths (e.g., memory CD4^+^ T cells with CDR3 AA length = 16, the Wilcoxon rank-sum test, *p*-value = 0.016).

To more formally assess the relative magnitude of RA-CDR3 scores across clinical and temporal groups, we performed Bayesian posterior comparisons using Markov Chain Monte Carlo (MCMC) sampling from the fitted models, accounting for variation in score distribution across different CDR3 AA lengths. At the cell level (**Fig. 3d**), at the sample from the time of onset of clinical RA, Converters showed higher RA-CDR3 scores when compared to their baseline sample, with a posterior probability of 93%. Even at baseline, Converters showed higher scores than Nonconverters (92%). In contrast, the probability of an increase from baseline to follow-up among Nonconverters was low (35%), suggesting a specific trajectory of autoreactive TCR accumulation in Converters. Similarly, sample-level analysis (**Supplementary Figure 5e**) corroborated these trends: Converters at follow-up had higher scores than both their own baseline (98%) and Nonconverters at follow-up (99%), and even at baseline, Converters scored higher than Nonconverters (98%).

These findings extend current knowledge by demonstrating that RA-CDR3 risk signals are enriched at baseline in individuals who will go on to develop RA and continue to expand modestly over time, supporting a model of early autoreactive T cell selection prior to clinical disease onset^34^.

### Induction of age-associated B cell signatures, altered somatic hypermutation, and reduced clonal diversity characterize preclinical B cell changes in RA Converters

To investigate the longitudinal behavior of B cell subsets and their relevance to RA development, we analyzed both CITE-seq and mass cytometry datasets for compositional, transcriptional, and clonal changes. UMAP visualization of B cells in the CITE-seq data revealed three major clusters corresponding to naive B, memory B, and double-negative B (DNB) cells (**Fig. 4a**). In terms of cell type frequency, no significant change was observed at either baseline or follow-up (**Supplementary Figure 7a-b**). Similarly, analysis of the mass cytometry dataset identified naïve and memory B cell clusters, but again showed no association with RA conversion status (**Supplementary Figure 7c-e**). An integrated analysis combining both datasets yielded consistent results, with no evidence of Converter-associated changes (**Supplementary Figure 7f-i**). Moreover, longitudinal comparison within the same individuals between baseline and follow-up samples revealed no significant changes in B cell subset frequencies (CITE-seq, **Supplementary Figure 7j**; mass cytometry, **Supplementary Figure 7k**).

**Fig. 4:**
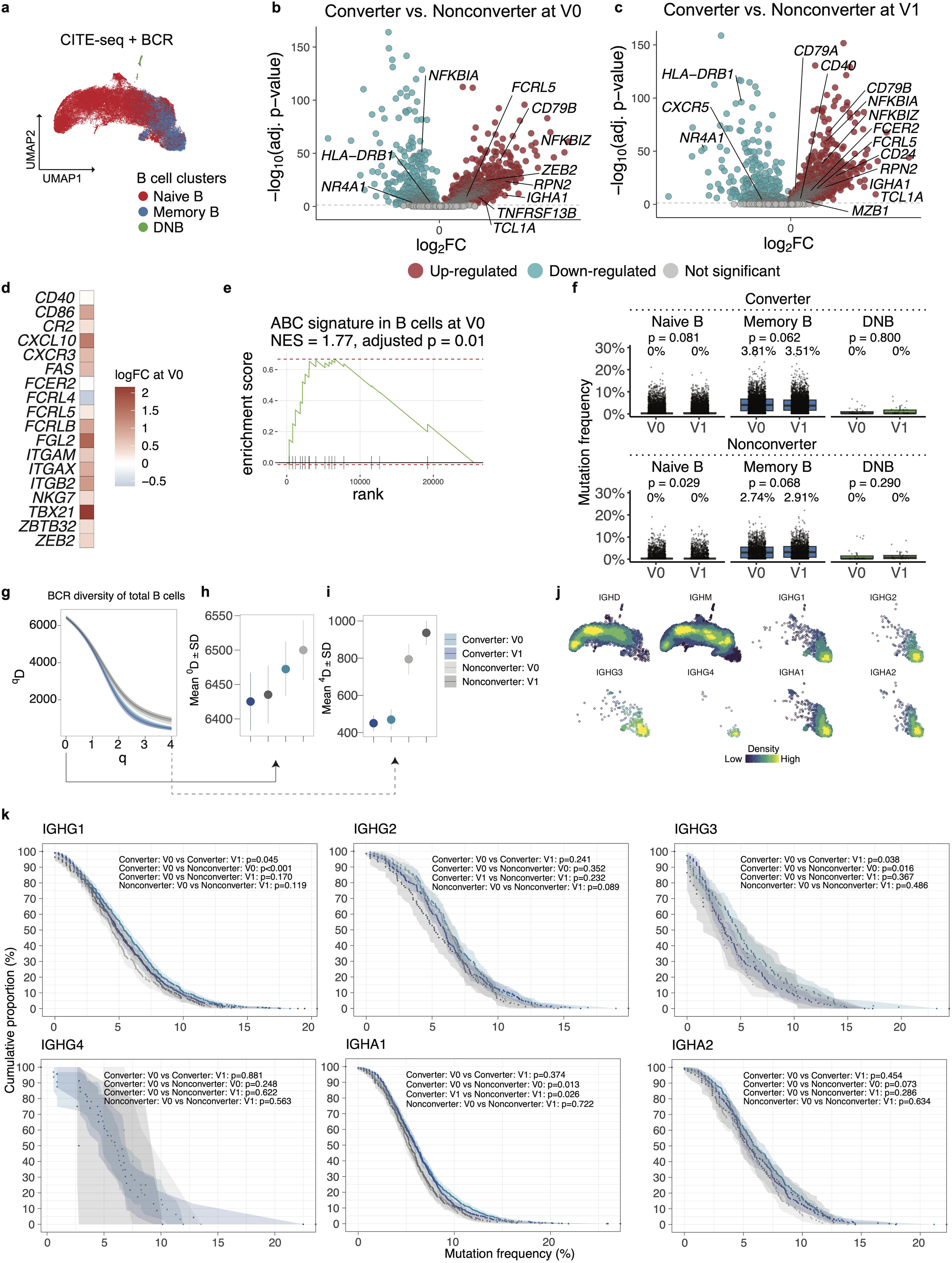
Multi-modal analysis of B cell subsets reveals transcriptional and clonal remodeling in RA Converters prior to disease onset. **a**, B cell clusters of CITE-seq dataset on UMAP. **b-c**. Differentially expressed genes in B cells between Converters and Nonconverters at baseline (V0, **b**) and post-onset/follow-up (V1, **c**). B-cell marker genes are annotated. **d**, Heatmap showing expression of representative age-associated B cell (ABC)-signature genes comparing RA Converters and Nonconverters at baseline. **e**, Normalized enrichment score (NES) by Gene Set Enrichment Analysis (GSEA) showing the enrichment of the ABC-signature in Converter at baseline. **f**, Somatic hypermutation frequencies (%) of BCR sequences in B cell subsets across time points in Converters and Nonconverters. P-values from Wilcoxon signed-rank tests compare V0 vs. V1 within each cell type and group. Mean mutation frequencies are indicated above each boxplot. **g**, BCR repertoire diversity by converter status and visit points. **h-i**, *D* zoomed at q=0 (**h**) and q=4 (**i**). **j**, Density plots of immunoglobulin isotypes on UMAP. **k**, Cumulative proportion (%) of BCR sequences with increasing mutation frequencies (x-axis) for specific Ig isotypes in Converters and Nonconverters, at baseline and post-onset/follow-up. Shaded areas represent confidence intervals. Statistical comparisons between time points and groups were performed using the Kolmogorov–Smirnov test.

Given the well-established role of B cells in RA pathogenesis^35,36^, including clonal expansion within the synovium^37^, we hypothesized that factors other than cell type frequency, such as transcriptional states beyond discrete cell types, may be associated with their contribution to disease development. To further dissect B cell transcriptional changes associated with RA development, we performed differential gene expression analysis between Converters and Nonconverters. At both baseline and follow-up, Converters displayed elevated expression of B BCR signaling components *CD79A* and *CD79B*^38,39^, alongside age-associated B cell (ABC) genes such as *FCRL5* and *ZEB2* (**Fig. 4b-c**), the latter known as a transcription factor that drives differentiation into ABCs^40^, as well as class-switched isotype *IGHA1*, which aligns with the mucosal origin hypothesis of RA^41,42^. A curated ABC-gene signature^40^ was elevated in Converters at baseline (**Fig. 4d-e**), although no longer reached statistical significance after RA onset (**Supplementary Figure 7l-m**). These results suggest that transcriptional changes associated with ABCs occur predominantly during a phase prior to the onset of clinical RA and potentially contribute to the transition to clinically apparent RA.

To further characterize B cell alterations during RA development, we performed BCR repertoire analysis. Converters exhibited a slight decrease over time, rather than increase, in somatic hypermutation (SHM) frequency after RA onset compared to prior to onset, although it was not significant (3.81% vs. 3.51%, the Wilcoxon rank-sum test, *p*-value=0.062) (**Fig. 4f**). This may reflect a shift in B cell activation toward extrafollicular or ectopic lymphoid pathways, possibly in situ, where true germinal centers are rare and affinity maturation is likely less efficient, as shown in published study SHM levels are lower in plasma cells from synovial fluid than those from peripheral blood^37^.

To quantitate clonal diversity, we computed Hill diversity indices (*D*) across varying *q* values. At both baseline and follow-up time points, B cells from Converters displayed lower overall diversity than Nonconverters consistently regardless of parameter *q* (**Fig. 4g-i**), suggesting dominance of clonally expanded populations, potentially expressing pathogenic anti-CCP autoantibodies.

It was clear that the overall distribution of immunoglobulin isotypes based on BCR sequencing was closely linked to cellular states by projection into the B-cell UMAP space (**Fig. 4j**), although no significant difference between Converters and Nonconverters over time between groups, or over time within groups (**Supplementary Figure 7n**). Importantly, however, evaluation of cumulative mutation frequency curves within immunoglobulin isotypes revealed elevated SHM levels in *IGHG1* and *IGHG3* among Converters compared to Nonconverters at baseline (Kolmogorov–Smirnov tests, *IGHG1* and *IGHG3, p*-value < 0.001 and *p*-value = 0.016, respectively; **Fig. 4k**). These mutation frequencies declined in Converters after RA onset (Kolmogorov–Smirnov tests, *IGHG1* and *IGHG3, p*-value = 0.045 and *p*-value = 0.038, respectively). In contrast, for *IGHA1*, while Converters also showed significantly higher SHM than Nonconverters at baseline (the Kolmogorov–Smirnov test, *p*-value = 0.013), there was no significant change over time (Converters at V0 and V1, the Kolmogorov–Smirnov test, *p*-value = 0.374). These isotype-specific patterns imply that class-switched B cells may undergo distinct trajectories in somatic hypermutation depending on their effector fate or site of activation, with IgG^+^ cells potentially more affected by extrafollicular responses than IgA^+^ cells.

Together with transcriptional and frequency-based findings, these results indicate that B cell remodeling in RA Converters involves induction of ABC signatures, and antigen-driven clonal selection, with many of these features emerging before clinical RA onset.

### NK cell subsets show distinct dynamics associated with RA conversion

We then focused on NK cell subsets using both mass cytometry and CITE-seq data. Here we prioritized the approach of analyzing separately mass cytometry (**Fig. 5a**), rather than CITE-seq and integration, considering limited overlap in protein panels between datasets and larger sample size in mass cytometry data. Mass cytometry analysis identified five NK cell subsets, annotated based on surface marker expression levels including CD56, CD16, CD57, CD137, and GZMB (**Fig. 5b-c**). Comparison of cell frequencies and GLMM analysis revealed that several subsets, particularly CD56^dim^CD16^+^CD57^+^, known as mature NK cells^43^, and CD56^dim^CD16^+^ expressing CD137, a costimulatory molecule that promotes proliferation and cytokine secretion^44,45^, were significantly expanded in RA Converters compared to Nonconverters at follow-up point (OR [95%CI] = 1.32 [1.08-1.61] for CD56^dim^CD16^+^CD57^+^ NK cells; OR [95%CI] = 1.14 [1.01-1.28] for CD56^dim^CD16^+^CD137^+^ NK cells), while a less cytotoxic subset, CD56^dim^CD16^+^GZMB^low^, were depleted in RA Converters at baseline (OR [95%CI] = 0.84 [0.73-0.99] for CD56^dim^CD16^+^GZMB^low^ NK cells) (**Fig. 5d-e**). Longitudinal analysis within each group revealed a significant decrease in CD56^bright^ NK cells in Nonconverters (the paired Wilcoxon signed-rank test, *p*-value = 0.0015), whereas other subsets remained largely stable between baseline and follow-up (**Supplementary Figure 9a**). In the analysis of CITE-seq data, no significant association was observed comparing Converters and Nonconverters at either baseline or follow-up (**Supplementary Figure 9c-f**).

**Fig. 5:**
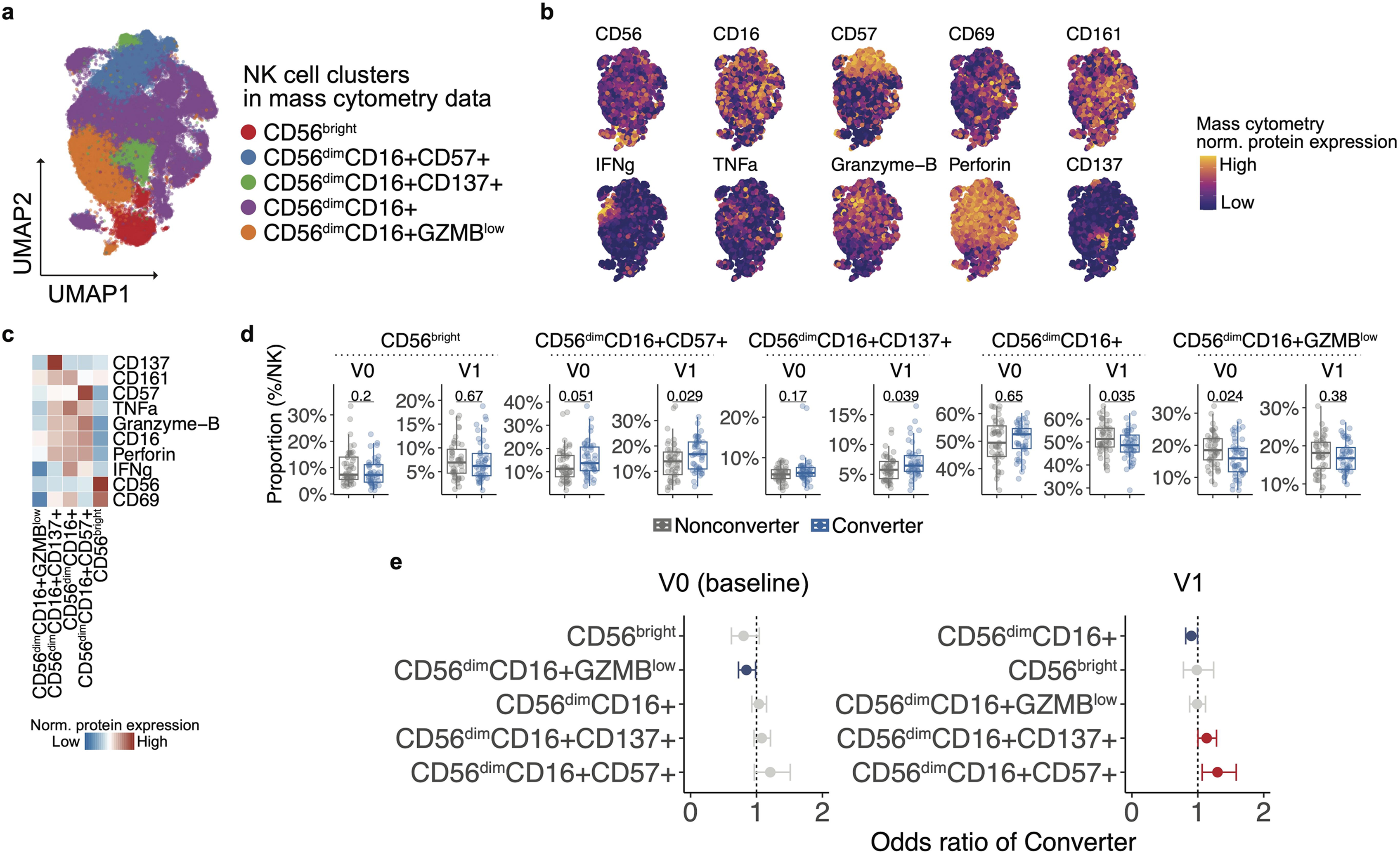
Characterization of NK cell subsets and association with RA conversion status. **a**, UMAP projection of NK cells from mass cytometry data. **b**, Surface protein expression levels of NK cell markers across the UMAP space. **c**, Heatmap showing average expression of key markers in each NK cell cluster. **d**, Box plots comparing the proportions of each NK subset (as a percentage of total NK cells) between Converters and Nonconverters at baseline (V0) and post-onset/follow-up (V1). P values from Wilcoxon rank-sum tests are shown above each comparison. **e**, GLMM analysis using mass cytometry NK cells comparing RA Converters and Nonconverters at each visit point, at baseline (V0, left) and post-onset/follow-up (V1). The model accounted for age, sex, batch and individuals as covariates. The odds ratio and 95% confidence intervals (CIs) are shown by error bars. CIs entirely above 1 are shown in red, indicating a positive association with RA conversion, while those entirely below 1 are shown in blue, indicating a negative association.

We next evaluated myeloid cells. CITE-seq analysis resolved five myeloid subsets including classical monocytes (cM), nonclassical monocytes (ncM), intermediate monocytes (intM), myeloid dendritic cells (mDCs), and plasmacytoid dendritic cells (pDCs) (**Supplementary Figure 9g-h**). GLMM revealed expansion of cM (OR [95%CI] = 1.37 [1.08-1.75] for cM) and depletion of mDCs in Converters at follow-up (OR [95%CI] = 0.75 [0.59-0.94] for mDCs) (**Supplementary Figure 9i-j**), but not significant in mass cytometry data analysis, where we identified four myeloid subsets: cM, ncM, mDC, and pDCs (**Supplementary Figure 9k-n**).

### Effect of HCQ

The analysis of the immunomodulatory effects of HCQ is detailed in the **Supplementary Text**. Briefly, while the StopRA trial concluded that HCQ did not prevent the onset of clinical RA, our detailed immunological analysis identified pro-inflammatory shifts associated with RA conversion—such as the expansion of Tph and memory CD4+ T cells, differentiation into memory B cells, and increases in specific NK cell subsets—were prominent only in the placebo group and were attenuated in the HCQ group. These findings suggest that while HCQ did not prevent clinical onset, it may partially modulate these immune pathways associated with disease progression.

### Chromatin accessibility profiling reveals cell-type-specific and conserved regulatory changes in individuals who developed RA

Because epigenomic states precede transcriptional changes, early alterations in chromatin accessibility could provide insights into the molecular mechanisms driving RA development. Therefore, we explored epigenomic alterations associated with progression to RA using scASAP-seq, which was available on a subset of 18 Converters and 8 Nonconverters (**Supplemental Table 1d**). As illustrated in the schematic (**Supplementary Figure. 10a**), we identified “Converter-related” peaks, that are regions of accessible chromatin enriched in Converters compared to Nonconverters, at both baseline and post-onset/follow-up (**Methods**). Here, we defined potential Converter-related peaks using nominal p-values < 0.05, given our limited statistical power due to the moderate sample size of scASAP-seq data to search across genomic regions.

After filtering in potential open regions in Converters (nominal *p*-value < 0.05 and effect size > 0), myeloid and NK cells contributed the largest number of Converter-related peaks at both baseline and follow-up (**Supplementary Figure 10b**). Converter-related peaks demonstrated significantly higher evolutionary conservation, as reflected by higher phastCons scores compared to other chromatin-accessible regions (**Supplementary Figure 10c**), supporting their functional relevance in RA pathogenesis. These Converter-related loci included known immunoregulatory functions and have been linked to autoimmune pathogenesis. Converter-specific accessibility signals in NK and myeloid cells were found near *STAT4* (**Supplementary Figure 10d**) and *NLRP3* (**Supplementary Figure 10e**), respectively. STAT4 is a key transcription factor required for IL-12–mediated cytotoxicity and IFN-γ production in both murine and human NK cells^46,47^. NLRP3 has been implicated in the hyperactivation of the inflammasome in myeloid cells, contributing to severe organ damage in experimental lupus^48^.

Collectively, these results demonstrate that epigenomic profiling of peripheral immune cells captures early and cell-type–specific regulatory shifts in individuals progressing to RA.

### Predictive modeling of RA onset using baseline immune and clinical features

Finally, we assessed whether clinical or immune features at baseline could predict future onset of RA. Two key questions were queried (**Fig. 6a**): (RQ1) whether conversion to RA was predictable at baseline using immune and clinical features, and (RQ2) whether these features were associated with the time to clinical RA onset in Converters. With future clinical applications in mind, we aimed to develop a simple and interpretable model by prioritizing features that can be readily measured in real-world settings, such as cell type frequencies that could ultimately be measurable by flow cytometry and standard clinical parameters, rather than a complex comprehensive framework. Accordingly, we built a composite predictive model that incorporated both prior knowledge of clinical variables and insights from our cellular analyses. Based on findings related to T cells, we used the frequency of cell types in the joint embedding space of CITE-seq and mass cytometry data as the cellular input. Specifically, the final model included the proportions of Tph, *GZMK*^+^*XCL1*^+^ CD8, and *GZMB*^+^*CD57*^+^ CD8^+^ T cells at baseline as cellular features, along with clinical variables such as age, sex, serum anti-CCP3 levels, RF-positivity, and dosage of shared epitope alleles in HLA loci.

**Fig. 6:**
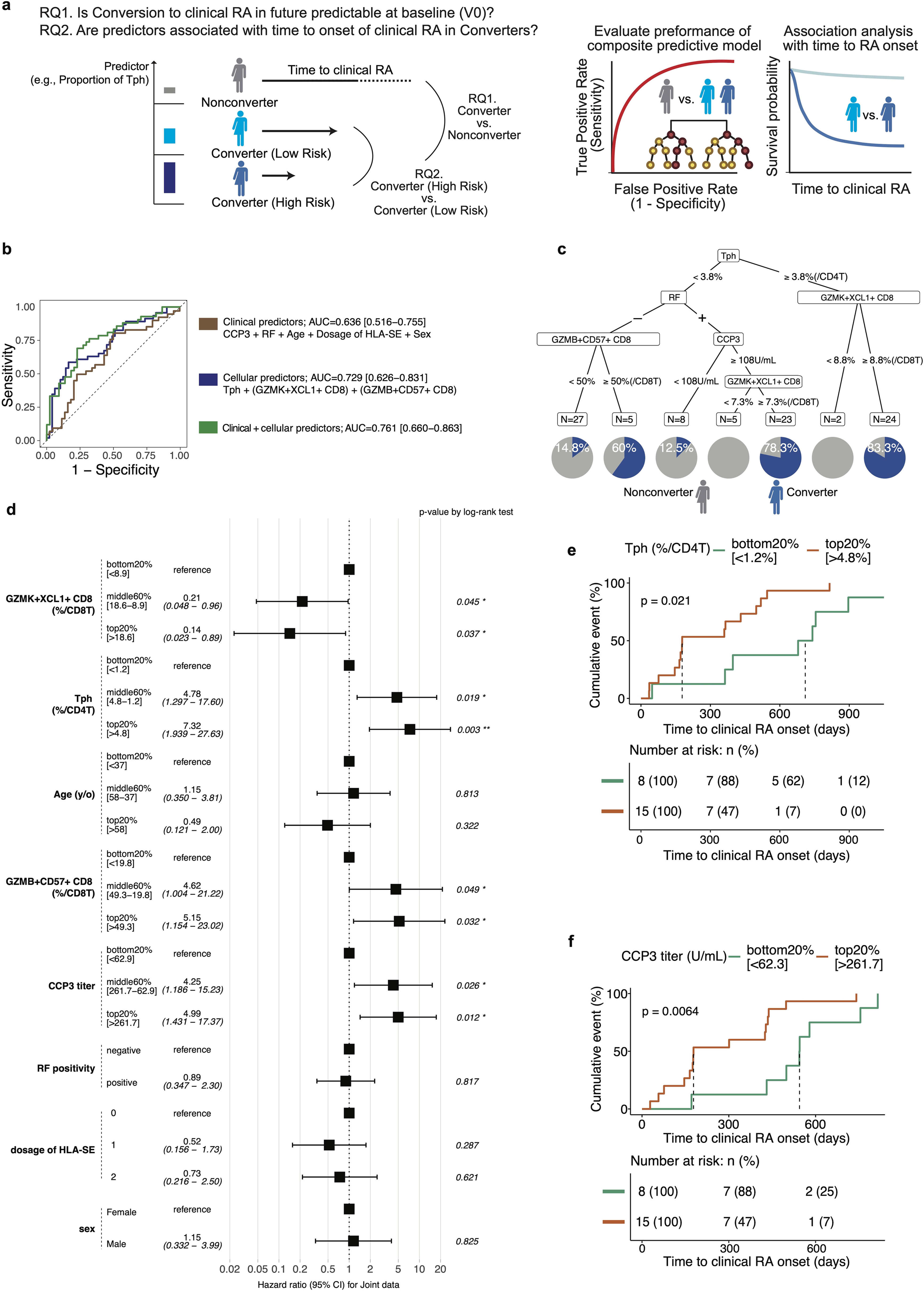
Predictive modeling of RA onset using baseline immune and clinical features. **a**, Schematic of the predictive modeling approach with research questions. **b**, Receiver operating characteristic (ROC) curves comparing the performance of models based on (1) Clinical predictors (brown): CCP3, RF, age, sex, and dosage of shared-epitope (SE) alleles in HLA loci, (2) Cellular predictors (blue): Tph, *GZMK*^+^*XCL1*^+^ CD8, and *GZMB*^+^*CD57*^+^ CD8^+^ T cells, and (3) Combined clinical + cellular predictors (green). **c**, Decision tree identifying risk strata for RA onset based on CCP3 levels, RFIgM positivity, Tph frequency (% of CD4^+^ T cells), and *GZMK*^+^*XCL1*^+^ CD8^+^ T cell frequencies (% of CD8^+^ T cells). Pie charts at terminal nodes indicate the proportion of Converters (blue) and Nonconverters (gray). **d**, Forest plot showing hazard ratios (HR) and 95% confidence intervals (CI) for time to RA onset among Converters, based on baseline immune and clinical features. Features are grouped by tertile or quantile bins, and p-values are from Cox proportional hazards models. **e-f,** Survival analysis of RA conversion stratified by Tph and CCP3 levels. Kaplan-Meier curves showing time to RA onset based on (**e**) top and bottom 20% of Tph proportions and (**f**) top and bottom 20% of CCP3 levels. P-values were calculated using log-rank tests.

To evaluate variable importance to classify Converter versus Nonconverters at baseline, we calculated mean decrease in Gini impurity in the random forest model (**Supplementary Figure 11a**). Tph cells and *GZMK*^+^*XCL1*^+^ CD8^+^ T cells ranked highest, followed by clinical variables, underscoring the importance of cellular biomarkers. Receiver operating characteristic (ROC) analysis demonstrated that immune cell-based predictors (AUC = 0.729 [0.626–0.831]) slightly outperformed clinical predictors (AUC = 0.636 [0.516–0.755]) in distinguishing Converters from Nonconverters (**Fig. 6b**). The combined model yielded the highest performance, with AUC of 0.761 [0.660–0.863]. To estimate the amount of risk explained by this model, a decision tree model identified subgroups with distinct probabilities of converting to RA, ranging as high as 83.3% based on these predictors (**Fig. 6c**).

We next focused on predicting time to clinical RA onset (treated as a continuous variable) specifically within Converters (RQ2). In this analysis, individuals were stratified into three groups based on the relative abundance of each feature: top 20%, bottom 20%, and middle 60%. A forest plot of hazard ratios (HRs) derived from multivariable Cox proportional hazards modeling revealed that individuals in the top 20% of Tph (≥4.8% of CD4^+^ T cells) and *GZMB*^+^*CD57*^+^ CD8^+^ T cells (≥49.3% of CD8^+^ T cells) were significantly associated with earlier RA onset compared to those in the bottom 20% (≤1.2% of CD4^+^ T cells and ≤19.8% of CD8^+^ T cells, respectively) (log-rank tests, *p*-value = 0.003 for Tph cells; *p*-value = 0.032 for *GZMB*^+^*CD57*^+^ CD8^+^ T cells) (**Fig. 6d**). Higher serum CCP3 levels (≥261.7 U/mL) also contributed significantly to prediction (the log-rank test, *p*-value = 0.012). Stratified Kaplan-Meier analysis confirmed that individuals in the top 20% for Tph frequency and CCP3 titer had significantly shorter times to RA onset (median: 178 days for Tph and 178 days for CCP3, respectively) compared to those in the bottom 20% (median: 710 days for Tph and 544 days for CCP3, respectively) (log-rank tests, *p*-value = 0.021 and *p*-value = 0.0064, respectively) (**Fig. 6e-f**). Despite being identified as significant in the Cox model, *GZMB*^+^*CD57*^+^ CD8^+^ T cells did not show a significant difference in Kaplan-Meier survival analysis (the log-rank test, *p*-value = 0.33) (**Supplementary Figure 11d**), suggesting their effect size may be context-dependent.

Together, these results demonstrate that combining immune cell subsets identified through single-cell multi-omic profiling with traditional clinical variables can enhance the ability to stratify preclinical RA risk and predict disease onset timing.

## Discussion

In this longitudinal single-cell multi-omic study of individuals at-risk for RA, we delineated early and persistent immune alterations that precede the onset of clinical RA. This prospective design that leverages the participants, data and biospecimens from a rigorously designed and executed formal clinical trial in RA prevention complements and expands prior cross-sectional and longitudinal studies of RA development^15–25^ by providing a distinct focus on the clonal and prognostic dimensions of RA progression. Specifically, we demonstrate that the observed expansions of Tph and *GZMK^+^*/*GZMB^+^* CD8^+^ T cells are characterized by clonal expansion. We also extended the analysis to include detailed BCR repertoire analysis (CDR3/SHM) and specific alterations in NK cell subsets, which have not yet been longitudinally characterized in the development of RA. Importantly, we translated these high-dimensional findings into a predictive model combining baseline cellular subsets and clinical factors to robustly classify individuals who progressed to RA and prognosticate disease timing. Together, these findings define early multi-omic immune signatures characterizing preclinical RA.

Tph cells have been implicated in seropositive RA as key drivers of extrafollicular B cell responses as we and others reported previously^2,15,17^. Extending those reports, we detected a significantly higher frequency of Tph cells in individuals who went on to develop RA compared to Nonconverters, even before any joints exhibited clinical synovitis. Mechanistically, Tph cells can home to inflamed sites and provide B-cell help via IL-21, CXCL13, and other mediators; our data raise the possibility that such interactions (for example, in lymph nodes or subclinical synovial infiltrates) are already active in the pre-RA stage. Furthermore, at the time point when clinical RA had developed, we further observed upregulation of a *CD96^high^* CD4⁺ T cell transcriptional signature. As for implications of this finding, the *CD96^high^*CD4^+^ T cells are regulated by the AHR/JUN axis, which suppresses CXCL13 and promotes IL-22 production^49^. Th22 cells have been shown to preferentially infiltrate RA synovium, where they promote osteoclastogenesis and bone destruction via IL-22–mediated signaling^50^. This highlights a potential immunological shift in CD4^+^ T cell states following RA onset.

In parallel with the Tph expansion, we found that Converters had increased populations of activated CD8^+^ T cells with distinctive phenotypes including *GZMK*^+^*XCL1*^+^ and *GZMB*^+^*CD57*^+^ subsets. These phenotypes are indicative of highly differentiated, chronically stimulated CD8^+^ T cells: *GZMK*^+^ cells often represent an effector memory subset with inflammatory potential^51–53^, whereas *GZMB*^+^*CD57*^+^ cells are typically terminally differentiated cytotoxic T cells that have undergone clonal expansion (CD57 marks replicative history^54^). Recent studies of patients with established RA have described clonally expanded CD8^+^ T cells expressing granzymes and cytotoxic molecules in both blood and synovium^51,55^. Our findings novelly suggest that these cells are present even at a prior stage where systemic autoimmunity is present yet clinical synovitis has not yet developed where the immune repertoire of Converters is skewed towards these effector CD8^+^ subsets. The concurrent presence of aberrant CD8^+^ T cell populations in parallel with helper T cells in Converters highlights a broad T cell dysregulation as a feature of RA development, involving multiple arms of the adaptive immune system.

Converters’ CD4^+^ T cells also had elevated CDR3 risk scores for RA, consistent with the idea that these individuals harbor T cell clones tuned to recognize joint-specific or citrullinated self-antigens. Indeed, increased T cell clonality in peripheral blood has been reported as individuals get closer to developing RA^34^￼. We speculate that the expanded T cell clones in Converters could be responding to the same antigens that drive autoantibody production (e.g. citrullinated peptides) or other concomitant triggers (such as microbiome^56^).

Our findings also demonstrate that, longitudinally, Converters accumulated B cells expressing ABC markers compared to Nonconverters. Functionally, this suggests a shift in Converters toward an extrafollicular B cell activation pathway, is likely driven by the expanded Tph cells which is also supported by prior published where Tph cells and ABCs have been shown to correlate and co-localize in autoimmune settings^57,58^. We also found that, in Converters, memory B cells’ Ig genes decreased in mutations (lower SHM load) over time, while SHM of Nonconverters’ memory B cells increased. This is an intriguing observation, given that high-affinity autoantibodies like anti-CCP are known to undergo extensive SHM during their development^59^, reflecting a germinal center-driven process, rather than extrafollicular sites, at the preclinical stage￼^60^. Our findings suggest that, during pre-RA stage, both germinal center (Tfh-dependent) and extrafollicular (Tph-dependent) modes of B cell activation may coexist, with a potential shift in dominance from a classical germinal center response that produces highly mutated, high-affinity antibody-secreting cells toward an extrafollicular pathway that gives rise to more polyclonal and less mutated B cell populations.

In the evaluation of epigenetic landscape, we identified dozens of potential differentially accessible regions in Converters versus Nonconverters. While previous studies have emphasized that RA risk SNPs are enriched in T cell enhancer regions and that established RA patients show distinct epigenomic signatures (e.g., altered methylation or chromatin marks) in T cells^61,62^, our data reveal that, in the preclinical stage, myeloid and NK cells contributed an even greater number of Converter-related chromatin accessibility peaks than T cells. One possibility is that T cells remain key epigenetic “hubs” in established RA, whereas pathogenesis during the pre-RA stage involves epigenetic dysregulation across multiple immune lineages, although T cell–associated changes become more pronounced once clinical symptoms manifest. Crucially, because our Converters had not yet received any immunologic treatments for RA and had no clinically-apparent synovitis, the epigenetic differences we observe likely represent cause or predisposition rather than consequence of disease or treatment.

We acknowledge several limitations of this study. First, the sample size, while one of the largest to date for a longitudinal single-cell study in the field of pre-clinical RA, was still limited in absolute numbers. RA conversion in high-risk cohorts is relatively infrequent, and our multi-omic analyses were constrained to those participants who had sufficient cells and samples across time. This might limit the statistical power to detect smaller-effect differences and could overfit some findings to this specific cohort. An external dataset for replication with the same measures was not available. Second, our study population consisted of individuals with anti-CCP positivity, essentially representing those at risk for seropositive RA. Our conclusions may not extrapolate to other at risk groups, such as first-degree relatives without antibodies, RF positivity without anti-CCP, those at-risk for seronegative RA, or people with subclinical imaging abnormalities but no antibodies. Third, we relied on peripheral blood sampling, which is a convenient and clinically relevant compartment but may not fully reflect the immune processes in target tissues like the synovium or lymph nodes. Additionally, some Nonconverters could still develop RA later, beyond our observation window; in that case, certain “Nonconverters” might have had early immune changes present that we misclassified as not being from a pre-RA state. This potential misclassification would, if anything, make our reported differences more conservative (diluting the Converter vs Nonconverter contrast).

In conclusion, by comprehensively profiling the immune system of individuals before and during their journey to RA, this study demonstrates that the path to arthritis is paved with identifiable and actionable immune changes. These insights lay the groundwork for transforming the care of at risk individuals through early identification and intervention, moving us closer to a future in which the development of RA can be foreseen and interdicted.

## Methods

### Subject recruitment and clinical data collection

Participants were enrolled in the NIH/NIAID-funded “Strategy to Prevent the Onset of Clinically-Apparent RA” (StopRA) trial^26^ (NCT02603146 and NIH designation ARA08), conducted in the United States from 2016 to 2022. The parent StopRA trial randomized a total of 144 individuals. This mechanistic study was approved by the sponsor (NIH/NIAID) and the Institutional Review Board at University of Colorado School of Medicine (protocol number COMIRB # 23-0490). For the StopRA trial, participants were enrolled who were positive for the anti-CCP3 antibody (Werfen, San Diego, California, USA) at a level ≥2 times the upper limit of normal and did not have clinical examination findings or historical evidence of inflammatory arthritis (IA) at baseline. All participants were randomized in a 1:1 ratio to receive hydroxychloroquine (HCQ) or placebo for one year, followed by up to two additional years of follow-up after cessation of treatment. The primary endpoint for the clinical trial was the development of ‘clinical RA’ that was defined by the 2010 American College of Rheumatology/European Alliance of Associations for Rheumatology (ACR/EULAR) classification criteria for RA^63^ with a score of ≥6, or a joint examination with RA-like synovitis with ≥1 bone erosion identified via x-ray imaging of the hands, wrists and feet. Evaluation for clinical RA was triggered by the presence on physical examination of ≥1 swollen joint consistent with RA-like synovitis. A secondary endpoint was termed ‘any IA’ and was defined as the development of ≥1 swollen joint consistent with IA with or without meeting the trial’s definition of clinical RA. Of the 142 participants in the StopRA modified intent-to-treat (mITT) population, 45 developed ‘clinical RA’ and 97 participants were designated as Nonconverters. For this mechanistic study, Converters (n=47) were designated as trial participants (n=45) who developed clinical RA at a research study visit and based on the trial’s formal definition for clinical RA, or were diagnosed with RA during the course of the study by a clinician outside of a trial visit (n=2), and Nonconverters were designated as those who did not develop clinical RA or any IA during the trial; specifically, participants (n=4) who developed IA not meeting the trial’s definition for clinical RA were excluded from this mechanistic study.

In the StopRA clinical trial, 144 participants were enrolled, and 142 were included in the primary modified Intent-to-Treat (mITT) analyses. Of these 142 participants, 45 developed clinical RA (21/69 [30.4%] % in the HCQ arm, and 24/73 [32.9%] in the placebo arm. The risk of clinical RA at 36 months was 0.336 with HCQ and 0.394 with placebo (difference -0.058; 95% confidence interval -0.336 to 0.220; p=0.52), indicating that 1 year of HCQ was not effective in reducing rates of clinical RA at 36 months.

For the mass cytometry studies, we analyzed a subset of the StopRA trial’s mITT population (**Supplemental Figure 1a**): 47 participants who developed clinical RA (Converters; 21 on HCQ, 26 on placebo), and 49 who did not develop clinical RA or IA (Nonconverters; 24 on HCQ, 25 on placebo) and who were frequency-matched to Converters by age, sex, and race.

For Converters, samples were selected based on adequate sample availability, and the earliest sample (V0) was primarily from the baseline visit with a follow-up sample selected from the time of onset of clinical RA (V1); in addition, for some Converters, an intermediate time point (V0.5) prior to clinical RA onset was also selected. For Nonconverters, samples were selected based on adequate sample availability, and the earliest sample (V0) was primarily from the baseline visit with a follow-up sample primarily from week 52 (V1). Notably, in Nonconverters, when possible, we selected a follow-up sample from a time point at which there was at least 1 year of clinical trial follow-up after that visit without incident clinical RA or IA. The rationale for this approach was to minimize effects in the Nonconverter follow-up samples from potential imminent clinical RA or IA development.

For the CITE-seq, B/T receptor sequencing and ASAP-seq analyses, we selected a subset of participants from those tested for mass cytometry based on sample availability. The sample time points (e.g., baseline, and incident clinical RA or follow-up) were the same as those selected for the mass cytometry assays; however, participants for CITE-seq, B/T cell receptor sequencing and ASAP-seq were selected only from the placebo arm in order to avoid confounding effects of HCQ use. The specific sample sets for each assay were as follows: for CITE-seq, n=24 Converters and n=10 Nonconverters; for T cell receptor sequencing n=24 Converters and n=9 Nonconverters; for B cell receptor sequencing, n=24 Converters and n=10 Nonconverters; for ASAP-seq, n=18 Converters and n=8 Nonconverters (**Supplemental Figure 1a**).

### Autoantibody and shared epitope testing

Anti-cyclic citrullinated peptide (anti-CCP) antibody testing was performed by enzyme linked immunosorbent assay (ELISA) (IgG CCP3, Quanta Lite, Werfen, San Diego, California USA). The level of positivity for this assay is ≥20 per the manufacturer’s suggestion; however, for the trial, the anti-CCP3 level for inclusion was ≥40 units which is ≥2 times the ULN. Rheumatoid factor immunoglobulin M (RF-IgM) testing was performed by ELISA (Quanta Lite, Werfen, San Diego, California USA). The level for positivity was established by a laboratory standard as a level present in <2% of controls without RA.

Anti-CCP and RF-IgM testing was performed at single laboratory (Exsera Biolabs, Inc., at the University of Colorado, Aurora, CO USA).

Human Leukocyte Allele (HLA) BRB1 testing was performed using real-time polymerase chain reaction to identify alleles considered to contain the ‘shared epitope’ which are sequences that are associated with risk for RA. For this study, the alleles considered to contain the shared epitope were: HLA DRB1*04:01, *04:04, *04:05, *04:08, *04:09, *04:10, *04:13, *01:01, *01:02, *10:00. Testing was performed at a single laboratory (ClinImmune Cell and Gene Therapy at the University of Colorado).

### Collection and processing of samples and preprocessing of single-cell multi-omic assays

See **Supplementary Notes**.

### Differential expression and gene set enrichment analyses

Differential gene expression (DE) analysis was performed using a custom Wilcoxon rank-sum test–based function (RunDEtest) in the SCP R package (v0.5.6)^64^. Only genes with a log2 fold change (log2FC) > 1 and Bonferroni adjusted p-value < 0.05 were considered statistically significant. To evaluate the enrichment of gene signatures across pseudobulk expression profiles, we performed gene set enrichment analysis (GSEA) using the fgseaMultilevel function from the fgsea R package^65^. Genes were ranked by average log2FC. For each gene list of signatures, see **Supplementary Notes**. From these gene sets for each signature, only genes that were detected in at least 5% of cells were included in the enrichment analysis. For each tested gene set, normalized enrichment score (NES) and Bonferroni adjusted p-values were reported, and enrichment plots were generated accordingly.

### Integration of CITE-seq and mass cytometry

To enable integrative analysis of protein expression across CITE-seq and mass cytometry (CyTOF) platforms, we employed the StabMap framework^66^ to generate a joint embedding space that preserves shared biological structure while accounting for modality-specific variation. Protein markers measured in each platform were mapped to corresponding gene symbols using curated dictionaries specific to CITE-seq and CyTOF panels. Overlapping 33 protein (LAMP1, CCR6, IL3RA, CD19, CD4, CD8A, ITGAX, FCGR3A, CD45RO, CD45RA, KLRB1, CCR4, IL2RA, CD27, B3GAT1, CXCR3, CXCR5, TIGIT, CD28, CD38, CD69, NCAM1, TRDV2, CD14, CD3E, MS4A1, HLA-DR, IGHD, IL7R, TNFRSF9, TNFRSF4, PDCD1, and ICOS) markers between the two modalities were retained for integration. This integration process was performed independently by broad cell types (T, B, myeloid, and NK cells). Normalized protein expression matrices were constructed separately for CITE-seq and CyTOF data. The resulting sparse matrices were stored in a named list and used as input to the stabMap function with CITE-seq specified as the reference modality. StabMap was configured to compute 20 components for both the reference and query modalities, and embeddings were scaled and centered prior to integration. Following integration, the imputed embeddings of all cells were obtained using the imputeEmbedding function, with a neighborhood size of five and CITE-seq as the reference. We applied UMAP to the joint StabMap embedding for visualization of modality-agnostic cellular structure. To propagate CITE-seq-derived cluster annotations into the shared embedding, we performed k-nearest neighbor (kNN) classification (k = 30) using the CITE-seq cluster labels. The trained kNN classifier assigned CITE-seq cluster identities to query cells in the mass cytometry dataset, allowing for cross-modality label transfer.

To evaluate the consistency of cell type classification across modalities, we assessed the correspondence between CITE-seq–based predicted clusters and manually annotated clusters from mass cytometry data. For each cell in mass cytometry data, we recorded both its original cluster assignment (mass cytometry-derived) and the corresponding predicted cluster label from CITE-seq. We constructed a contingency table of predicted CITE-seq clusters versus original mass cytometry clusters. To quantify the overlap, we computed the match rate by normalizing each row of the contingency table to obtain the proportion of mass cytometry cells from each predicted cluster that matched each mass cytometry-annotated cluster.

### Repertoire analysis

In order to assess the expansion of clones for TCR and BCR, gene segment (V, D, J) usage frequency, Isotype assignment, BCR mutation frequency and somatic hypermutation (SHM) analysis, we implemented Change-O^67^ and scRepertorie (v1.12.0)^68^. Clonal diversity is calculated using the generalized diversity index (Hill numbers)^30^.

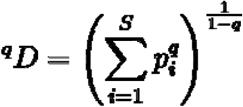

This method quantifies diversity as a smooth function (D) of a single parameter q. As the parameter q increases from 0 to + ∞ the diversity index (D) depends less on rare clones and more on common (abundant) ones, thus encompassing a range of definitions that can be visualized as a single curve. Large Hill Numbers are generally interpreted as high diversity in clonal populations.

To identify public TCRs with known antigens we focused on TCR β-chain (TRB) sequences. We curated a reference database of public TCRs from VDJdb (retrieved at 2023-10- 17), filtering for TRB chains. Each public TCR was represented as a combination of the CDR3 AA sequence and the V gene. TCRs from our dataset were similarly processed b concatenating the CDR3 AA sequence and the annotated V gene. We then identified public TCRs by performing an exact match between these sequences and the reference public TCR database.

### Calculation of RA-associated CDR3 risk scores for TCR

To quantify the autoimmune potential of individual T cell receptors, we computed CDR3 risk scores using position-specific effect sizes previously reported for RA by the cdr3-QTL framework^33^ (downloaded from https://github.com/immunogenomics/cdr3-QTL). Specifically, we used the dataset HIP_rascore_aausage_normbetas_v1.RData, which contains normalized beta coefficients representing RA effect sizes for individual AAs at specific CDR3β positions across different CDR3 lengths. For each TCR β-chain CDR3 AA sequence, we calculated a score by summing the corresponding effect sizes of AAs at each position, provided the length of the CDR3 sequence was between 12 and 17 AAs. Sequences outside this range were assigned a score of zero.

We visualized the distribution of risk scores and evaluated their associations with RA progression status (“Converter” vs. “Nonconverter”) and visit time point (V0 or V1). We further fitted a zero-inflated Gaussian model using glmmTMB to account for excess zero values in CDR3 scores, incorporating the AA length of CDR3β as a zero-inflation term. Estimated marginal means and pairwise comparisons were evaluated using the emmeans^69^ package to test directional hypotheses regarding score differences across groups (e.g., “Converter: V1 > Converter: V0”) using one-sided tests.

To further evaluate the differences in RA-associated CDR3 risk scores across clinical groups and time points, we implemented Bayesian hierarchical modeling using the brms R package^70^. After filtering in T cells with the length of TCR β-chain CDR3 AA between 12 and 17, two separate models were fit: a cell-level model using all individual TCRs and a sample-level model using mean scores aggregated per sample, visit, and CDR3 length. In both models, the outcome variable was the CDR3 risk score, and the main predictor was the interaction term between visit (V0 or V1) and RA progression status (“Converter” or “Nonconverter”) assuming a Gaussian likelihood. We included AA length as a random effect to account for variation in score distribution across different CDR3 lengths. For the sample-level model, scores were first averaged within each sample. Weakly informative priors were used for fixed effects (normal(0,1)) and standard deviations of random effects (Cauchy(0,1)). Posterior distributions were estimated using four Markov Chain Monte Carlo chains, each run for 2,000 iterations. For pairwise group comparisons, we computed posterior differences between group-specific estimates and evaluated one-sided posterior probabilities that one group had higher scores than another (e.g., P(Converter:V1 > Converter:V0)).

### Statistical models for Converter association tests

To assess the association between cell population frequencies and clinical outcome (Converter versus Nonconverter) at each visit point, we applied mixed-effects logistic regression models using the Mixed-effects Association of Single Cells (MASC) framework^71^. We implemented these models separately for the CITE-seq dataset, the mass cytometry dataset, and the integrated dataset derived from both platforms. In our analyses, we performed MASC as follows:

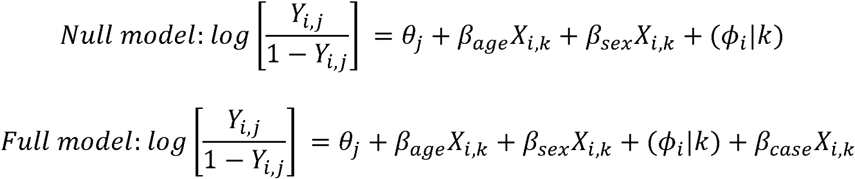

Here, *Y_i,j_* is the odds of cell *i* belonging to cluster *j*, θ*_j_* is the intercept for cluster *j*, β*_age_* and β*_sex_* indicate the fixed-effect of age and sex for cell *i* from k^th^ sample, respectively; (L*_i_* |*k*) is the random effect for cell *i* from k^th^ sample, β*_case_* indicates the effect of k^th^ sample’s RA clinical outcome status (Converter or Nonconverter). The model fitting was performed using the nloptwrap optimizer and Laplace approximation with adaptive Gauss–Hermite quadrature (nAGQ = 1). For the CITE-seq data, we included batch as a random effect to account for technical variation across sample processing batches. In the mass cytometry dataset, as with the CITE-seq model, batch was included as a random effect. For the integrated dataset, which combined CITE-seq and mass cytometry using the StabMap framework, we utilized predicted cell subtypes inferred from the shared embedding space as the cluster variable. The model included additional covariates to account for study design, such as the treatment arm for HCQ, which was incorporated as a fixed effect along with age and gender. A hierarchical random effects structure was introduced to account for nested sources of variation, including dataset, batch nested within dataset, and participant ID nested within batch and dataset. For each gene–peak pair, a likelihood ratio test (LRT) was performed to compare the full model (including the RA conversion status variable) against a null model (excluding this variable). We presented our results from MASC by odds ratio with an error bar indicating 95% confidence intervals for each cluster.

### Chromatin accessibility analysis

We investigated the chromatin accessibility profiles associated with RA conversion status using scATAC-seq data derived from the ASAP-seq platform. To characterize chromatin regulatory features linked to conversion to RA (Converter vs. Nonconverter), we constructed a gene-peak matrix based on peaks called in major immune cell subsets (T, B, myeloid, and NK cells), then intersected these peaks with proximity-based gene annotations using a ±500 kb window centered on each gene. We removed peaks of genes with bottom 75% fragment coverages. For statistical testing, a pseudobulk strategy was used within each major immune cell subset and each visit time point (V0, V1). Peak accessibility was binarized (accessible vs. not accessible). For sparsely accessible peaks (<5% accessibility), models were not evaluated due to insufficient data. Association with RA conversion status was tested using generalized linear models with binomial regression as follows;

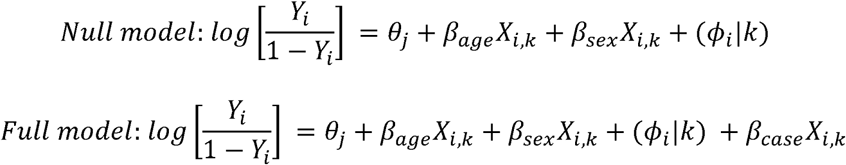

Where *Y_i_* is the odds of cell *i* for the binary peak accessibility, θ*_j_* is the intercept, β*_age_* and β*_sex_* indicate the fixed-effect of age and sex for cell *i* from k^th^ sample, respectively; (L*_i_* |*k*) is the random effect for cell *i* from k^th^ sample, and β*_case_* indicates the effect of k^th^ sample’s RA clinical outcome status (Converter or Nonconverter). For each gene–peak pair, a likelihood ratio test (LRT) was performed to compare the full model (including the RA conversion status variable) against a null model (excluding this variable). We filtered for potential peaks using the following thresholds by time points: nominal LRT p-value < 0.05 and regression coefficient β*_case_* > 0 for Converter enrichment.

To evaluate the evolutionary conservation of genomic regions with increased chromatin accessibility in RA Converters, we calculated conservation scores for Converter-related and background peaks using the phastCons100way.UCSC.hg38 track^72^, which represents conservation across 100 vertebrate species. Converter-related peaks (foreground) and all other peaks (background) were each scored based on overlap with phastCons elements.

### Predictive modeling and survival analysis

To identify predictors of progression to RA, we developed machine learning models using baseline immune profiling data, including annotated immune cell cluster proportions from CITE-seq and mass cytometry (cellular variables), in combination with clinical metadata. Data preprocessing was performed separately for each modality. For cellular variables, we included frequency of Tph, *GZMK^+^XCL1^+^* CD8, and *GZMB^+^CD57^+^*CD8 as predictors based on their enrichment in Converters at baseline. Clinical features included age, sex, anti-CCP3 antibody titer, RF-IgM positivity, and dosage of HLA-SE measured by PCR-SSP (polymerase chain reaction with sequence-specific primers) method. We calculated feature importance by random forest models to classify Converters vs. Nonconverters. Performance of composite predictive models which included clinical variables, cellular variables, and both were evaluated based on area under the receiver operating characteristic curve (AUROC). To visualize key immune and clinical predictors of RA conversion in an interpretable format, we constructed a classification decision tree model using the rpart package in R. Candidate features included anti-CCP3 antibody titer, RF-IgM positivity, and frequencies of Tph cells, *GZMK*^+^*XCL1*^+^ CD8^+^ T cells, and *GZMB*^+^*CD57*^+^ CD8^+^ T cells). The classification tree was fitted using the Gini index as the splitting criterion, with class priors set equally (0.5 for both classes). Model complexity was controlled by setting the maximum tree depth to 4, minimum bucket size to 1, and complexity parameter (cp) to 0, allowing the full tree to be grown for visualization purposes.

For time-to-event analysis, we applied Cox proportional hazards models to assess the association of baseline immune features with time to RA development. We also evaluated multivariate models adjusted for clinical covariates. Hazard ratios and 95% confidence intervals were reported. Kaplan–Meier survival curves were generated for representative features stratified by top 20% or bottom 20%, and statistical differences were evaluated using the log-rank test.

### Statistics

Differential gene expression analyses were conducted using the Wilcoxon rank-sum test (two-sided). For volcano plots, adjusted p-values were computed using the Benjamini–Hochberg method to correct for multiple testing. All statistical tests are noted in the corresponding figure legends and tables. A p-value < 0.05 was considered statistically significant unless otherwise specified.

## Data availability

All raw and processed data have been deposited in the Immport (http://XXX) under accession code XXX. Supplemental material for this paper is available online.

## Code availability

All source code to reproduce the analyses will be available on GitHub (https://github.com/fanzhanglab/Longitudinal_preRA) and Zenodo (https://doi.org/XXX).

## Data Availability

All data produced in the present study will be available upon acceptance.

## Acknowledgments

This research was performed as part of the Autoimmunity Centers of Excellence (ACE), and was funded by the National Institute of Allergy and Infectious Disease (NIAID) of the National Institutes of Health (NIH) (UM1 AI110503, U19 AI110483). In addition, this work was supported by the NIAMS R01-AR085156, an Arthritis National Research Foundation award, an Arthritis Foundation grant, and the University of Colorado School of Medicine Translational Research Scholars Program award (to FZ); NIAMS R01-AR080659, NIAMS R01-AR077607, NIAMS P30-AR070253, and NIAMS P30-AR079369 (to MLF, VMH and KDD); NIAMS P30AR073750, NIAID UM1AI144292, NIAID U01AI1176244 and NIGMS U54GM104938 (JAJ, JMG). This work was also supported by the Uehara Memorial Foundation Postdoctoral Fellowship, a Grant-in-Aid for Japan Society for the Promotion of Science Overseas Research Fellows, the Mochida Memorial Foundation for Medical and Pharmaceutical Research, and NIH/NCATS Colorado Clinical and Translational Science Awards grant UM1-TR004399 (to JI). We thank Wade deJager and Susan Macwana for their insights and support with sample processing.

## Author contributions

JAS, KDD, VMH, and MLF recruited patients, obtained samples, and curated clinical data. AB, MS, CW, TS, JAJ, and JMG contributed to sample procurement and processing, and to the design and implementation of the sample preparation, cell sorting, and data generation pipelines. JI led the computational and statistical analyses with input from LV, AB, MS. JI wrote the initial draft with guidance from FZ. FZ provided primary supervision of the computational work and co-supervised the study together with JMG, KD, and VMH. All authors participated in editing the final manuscript.

## References

1. Gravallese, E. M. & Firestein, G. S. Rheumatoid Arthritis - Common Origins, Divergent Mechanisms. N Engl J Med 388, 529–542 (2023).

2. Deane, K. D. et al. Therapeutic interception in individuals at risk of rheumatoid arthritis to prevent clinically impactful disease. Ann Rheum Dis 84, 14–28 (2025).

3. Frisell, T. et al. Familial risks and heritability of rheumatoid arthritis: role of rheumatoid factor/anti-citrullinated protein antibody status, number and type of affected relatives, sex, and age. Arthritis Rheum 65, 2773–2782 (2013).

4. Nielen, M. M. J. et al. Specific autoantibodies precede the symptoms of rheumatoid arthritis: a study of serial measurements in blood donors. Arthritis Rheum 50, 380–386 (2004).

5. Majka, D. S. et al. Duration of preclinical rheumatoid arthritis-related autoantibody positivity increases in subjects with older age at time of disease diagnosis. Ann Rheum Dis 67, 801–807 (2008).

6. van Steenbergen, H. W., da Silva, J. A. P., Huizinga, T. W. J. & van der Helm-van Mil, A. H. M. Preventing progression from arthralgia to arthritis: targeting the right patients. Nat Rev Rheumatol 14, 32–41 (2018).

7. Kokkonen, H. et al. Up-regulation of cytokines and chemokines predates the onset of rheumatoid arthritis. Arthritis Rheum 62, 383–391 (2010).

8. Deane, K. D. et al. The number of elevated cytokines and chemokines in preclinical seropositive rheumatoid arthritis predicts time to diagnosis in an age-dependent manner. Arthritis Rheum 62, 3161–3172 (2010).

9. Chang, H.-H., et al. A molecular signature of preclinical rheumatoid arthritis triggered by dysregulated PTPN22. JCI Insight 1, e90045 (2016).

10. Kelmenson, L. B. et al. Timing of Elevations of Autoantibody Isotypes Prior to Diagnosis of Rheumatoid Arthritis. Arthritis Rheumatol 72, 251–261 (2020).

11. Deane, K. D., Norris, J. M. & Holers, V. M. Preclinical rheumatoid arthritis: identification, evaluation, and future directions for investigation. Rheum Dis Clin North Am 36, 213–241 (2010).

12. Aho, K. et al. Antifilaggrin antibodies within ‘normal’ range predict rheumatoid arthritis in a linear fashion. J Rheumatol 27, 2743–2746 (2000).

13. Hemminki, K., Li, X., Sundquist, J. & Sundquist, K. Familial associations of rheumatoid arthritis with autoimmune diseases and related conditions. Arthritis Rheum 60, 661–668 (2009).

14. Anioke, I. et al. Lymphocyte subset phenotyping for the prediction of progression to inflammatory arthritis in anti-citrullinated-peptide antibody-positive at-risk individuals. Rheumatology (Oxford) 63, 1720–1732 (2024).

15. Inamo, J. et al. Deep immunophenotyping reveals circulating activated lymphocytes in individuals at risk for rheumatoid arthritis. J Clin Invest 135, (2025).

16. James, E. A. et al. Multifaceted immune dysregulation characterizes individuals at-risk for rheumatoid arthritis. Nat Commun 14, 7637 (2023).

17. Takada, H. et al. Expansion of HLA-DR Positive Peripheral Helper T and Naive B Cells in Anticitrullinated Protein Antibody-Positive Individuals At Risk for Rheumatoid Arthritis. Arthritis Rheumatol 76, 1023–1035 (2024).

18. Ponchel, F. et al. T-cell subset abnormalities predict progression along the Inflammatory Arthritis disease continuum: implications for management. Sci Rep 10, 3669 (2020).

19. Hunt, L. et al. T cell subsets: an immunological biomarker to predict progression to clinical arthritis in ACPA-positive individuals. Ann Rheum Dis 75, 1884–1889 (2016).

20. Lübbers, J. et al. Changes in peripheral blood lymphocyte subsets during arthritis development in arthralgia patients. Arthritis Res Ther 18, 205 (2016).

21. Chalan, P., Bijzet, J., Kroesen, B.-J., Boots, A. M. H. & Brouwer, E. Altered Natural Killer Cell Subsets in Seropositive Arthralgia and Early Rheumatoid Arthritis Are Associated with Autoantibody Status. J Rheumatol 43, 1008–1016 (2016).

22. Anang, D. C. et al. Increased Frequency of CD4 Follicular Helper T and CD8 Follicular T Cells in Human Lymph Node Biopsies during the Earliest Stages of Rheumatoid Arthritis. Cells 11, (2022).

23. Ramwadhdoebe, T. H. et al. Lymph node biopsy analysis reveals an altered immunoregulatory balance already during the at-risk phase of autoantibody positive rheumatoid arthritis. Eur J Immunol 46, 2812–2821 (2016).

24. Ramwadhdoebe, T. H. et al. Human lymph-node CD8(+) T cells display an altered phenotype during systemic autoimmunity. Clin Transl Immunology 5, e67 (2016).

25. He, Z. et al. Progression to rheumatoid arthritis in at-risk individuals is defined by systemic inflammation and by T and B cell dysregulation. Sci Transl Med 17, eadt7214 (2025).

26. Deane, K. D. et al. A phase 2 trial of hydroxychloroquine in individuals at risk for rheumatoid arthritis. Arthritis Rheumatol (2025) doi:10.1002/art.43366.

27. ClinicalTrials.gov. https://clinicaltrials.gov/study/NCT02603146.

28. Mimitou, E. P. et al. Scalable, multimodal profiling of chromatin accessibility, gene expression and protein levels in single cells. Nat Biotechnol 39, 1246–1258 (2021).

29. Zhang, F. et al. Defining inflammatory cell states in rheumatoid arthritis joint synovial tissues by integrating single-cell transcriptomics and mass cytometry. Nature Immunology 20, 928–942 (2019).

30. Hill, M. O. Diversity and evenness: A unifying notation and its consequences. Ecology 54, 427–432 (1973).

31. Ishigaki, K. et al. Multi-ancestry genome-wide association analyses identify novel genetic mechanisms in rheumatoid arthritis. Nat Genet 54, 1640–1651 (2022).

32. Dendrou, C. A., Petersen, J., Rossjohn, J. & Fugger, L. HLA variation and disease. Nature Reviews Immunology 18, 325–339 (2018).

33. Ishigaki, K. et al. HLA autoimmune risk alleles restrict the hypervariable region of T cell receptors. Nat Genet 54, 393–402 (2022).

34. Lamacchia, C. et al. Detection of circulating highly expanded T-cell clones in at-risk individuals for rheumatoid arthritis before the clinical onset of the disease. Rheumatology (Oxford) 60, 3451–3460 (2021).

35. Scherer, H. U., van der Woude, D. & Toes, R. E. M. From risk to chronicity: evolution of autoreactive B cell and antibody responses in rheumatoid arthritis. Nature Reviews Rheumatology 18, 371–383 (2022).

36. Wu, F. et al. B Cells in Rheumatoid Arthritis：Pathogenic Mechanisms and Treatment Prospects. Front Immunol 12, 750753 (2021).

37. Dunlap, G. et al. Clonal associations between lymphocyte subsets and functional states in rheumatoid arthritis synovium. Nature Communications 15, 1–21 (2024).

38. Pelanda, R., Braun, U., Hobeika, E., Nussenzweig, M. C. & Reth, M. B cell progenitors are arrested in maturation but have intact VDJ recombination in the absence of Ig-alpha and Ig-beta. J Immunol 169, 865–872 (2002).

39. Gong, S. & Nussenzweig, M. C. Regulation of an early developmental checkpoint in the B cell pathway by Ig beta. Science 272, 411–414 (1996).

40. Dai, D. et al. The transcription factor ZEB2 drives the formation of age-associated B cells. Science 383, 413–421 (2024).

41. Holers, V. M. et al. Rheumatoid arthritis and the mucosal origins hypothesis: protection turns to destruction. Nat Rev Rheumatol 14, 542–557 (2018).

42. Holers, V. M. et al. Distinct mucosal endotypes as initiators and drivers of rheumatoid arthritis. Nat Rev Rheumatol 20, 601–613 (2024).

43. Lopez-Vergès, S. et al. CD57 defines a functionally distinct population of mature NK cells in the human CD56dimCD16+ NK-cell subset. Blood 116, 3865–3874 (2010).

44. Ochoa, M. C. et al. Daratumumab in combination with urelumab to potentiate anti-myeloma activity in lymphocyte-deficient mice reconstituted with human NK cells. Oncoimmunology 8, 1599636 (2019).

45. Wilcox, R. A., Tamada, K., Strome, S. E. & Chen, L. Signaling through NK cell-associated CD137 promotes both helper function for CD8+ cytolytic T cells and responsiveness to IL-2 but not cytolytic activity. J Immunol 169, 4230–4236 (2002).

46. Townsend, M. J. et al. T-bet regulates the terminal maturation and homeostasis of NK and Valpha14i NKT cells. Immunity 20, 477–494 (2004).

47. Wang, K. S., Frank, D. A. & Ritz, J. Interleukin-2 enhances the response of natural killer cells to interleukin-12 through up-regulation of the interleukin-12 receptor and STAT4. Blood 95, 3183–3190 (2000).

48. Lu, A. et al. Hyperactivation of the NLRP3 Inflammasome in Myeloid Cells Leads to Severe Organ Damage in Experimental Lupus. J Immunol 198, 1119–1129 (2017).

49. Law, C. et al. Interferon subverts an AHR–JUN axis to promote CXCL13+ T cells in lupus. Nature 631, 857–866 (2024).

50. Miyazaki, Y. et al. Th22 Cells Promote Osteoclast Differentiation Production of IL-22 in Rheumatoid Arthritis. Front Immunol 9, 2901 (2018).

51. Jonsson, A. H. et al. Granzyme K CD8 T cells form a core population in inflamed human tissue. Sci Transl Med 14, eabo0686 (2022).

52. Donado, C. A. et al. Granzyme K activates the entire complement cascade. Nature 1–11 (2025).

53. Lan, F. et al. GZMK-expressing CD8+ T cells promote recurrent airway inflammatory diseases. Nature 638, 490–498 (2025).

54. Brenchley, J. M. et al. Expression of CD57 defines replicative senescence and antigen-induced apoptotic death of CD8+ T cells. Blood 101, 2711–2720 (2003).

55. Moon, J.-S. et al. Cytotoxic CD8 T cells target citrullinated antigens in rheumatoid arthritis. Nat Commun 14, 319 (2023).

56. Seymour, B. J. et al. Microbiota-dependent indole production stimulates the development of collagen-induced arthritis in mice. J Clin Invest 134, (2023).

57. Bocharnikov, A. V., et al. PD-1hiCXCR5- T peripheral helper cells promote B cell responses in lupus via MAF and IL-21. JCI Insight 4, (2019).

58. Zhang, F. et al. Deconstruction of rheumatoid arthritis synovium defines inflammatory subtypes. Nature 623, 616–624 (2023).

59. Vergroesen, R. D. et al. B-cell receptor sequencing of anti-citrullinated protein antibody (ACPA) IgG-expressing B cells indicates a selective advantage for the introduction of - glycosylation sites during somatic hypermutation. Ann Rheum Dis 77, 956–958 (2018).

60. Elsner, R. A. & Shlomchik, M. J. Germinal Center and Extrafollicular B Cell Responses in Vaccination, Immunity, and Autoimmunity. Immunity 53, 1136–1150 (2020).

61. Vahedi, G. et al. Super-enhancers delineate disease-associated regulatory nodes in T cells. Nature 520, 558–562 (2015).

62. Yang, J. et al. Analysis of chromatin organization and gene expression in T cells identifies functional genes for rheumatoid arthritis. Nat Commun 11, 4402 (2020).

63. Aletaha, D. et al. 2010 Rheumatoid arthritis classification criteria: an American College of Rheumatology/European League Against Rheumatism collaborative initiative. Arthritis Rheum 62, 2569–2581 (2010).

64. Website. https://github.com/zhanghao-njmu/SCP.

65. Korotkevich, G. et al. Fast gene set enrichment analysis. bioRxiv (2016) doi:10.1101/060012.

66. Ghazanfar, S., Guibentif, C. & Marioni, J. C. Stabilized mosaic single-cell data integration using unshared features. Nature Biotechnology 42, 284–292 (2023).

67. Gupta, N. T. et al. Change-O: a toolkit for analyzing large-scale B cell immunoglobulin repertoire sequencing data. Bioinformatics 31, 3356–3358 (2015).

68. Borcherding, N., Bormann, N. L. & Kraus, G. scRepertoire: An R-based toolkit for single-cell immune receptor analysis. F1000Res. 9, 47 (2020).

69. Estimated Marginal Means, aka Least-Squares Means. https://rvlenth.github.io/emmeans/.

70. Bürkner, P.-C. Brms: An R package for Bayesian multilevel models using Stan. J. Stat. Softw. 80, (2017).

71. Fonseka, C. Y. et al. Mixed-effects association of single cells identifies an expanded effector CD4 T cell subset in rheumatoid arthritis. Sci Transl Med 10, (2018).

72. Castelo, R. phastCons100way.UCSC.hg38. (Bioconductor, 2017). doi:10.18129/B9.BIOC.PHASTCONS100WAY.UCSC.HG38.

