## Supplementary Notes and Figures for "Longitudinal peripheral blood multi-omic profiling in seropositive individuals identifies immune endotypes and predictive models for future rheumatoid arthritis conversion"

**Inamo et al.**

**Table of Contents:**


**Page 3 Supplementary Text**

**Page 9 Supplementary Notes**

**Page 16 Supplementary References**

**Page 18 Supplementary Figures**


**Supplementary Tables are provided by a separate excel file.**

### Supplementary Text

#### Summary of Methods and Findings

| Analysis Area | Methods Used | Key Findings |
|---|---|---|
| T Cell Profiling | • CITE-seq & Mass Cytometry Integration<br>• Generalized Linear Mixed Models (GLMMs)<br>• Differential Gene Expression (DEG) & Gene Set Enrichment Analysis (GSEA) | • Individuals who later developed RA ("Converters") had expansions of peripheral helper T (Tph) cells and cytotoxic *GZMK*$^{+}$ & *GZMB*$^{+}$ CD8$^{+}$ T cells at baseline, before clinical onset.<br>• A Tph-related transcriptional signature was significantly enriched in the CD4$^{+}$ T cells of Converters at baseline. |
| T Cell Receptor (TCR) Analysis | • Single-cell TCR sequencing<br>• Clonal diversity analysis (Hill diversity)<br>• RA-associated CDR3 risk score calculation and modeling | • In Converters, CD8$^{+}$ T cells underwent clonal narrowing after RA onset, while CD4$^{+}$ T cells maintained broad repertoire diversity.<br>• CD4$^{+}$ T cells in Converters had significantly higher RA-associated TCR risk scores at baseline compared to Nonconverters. |
| B Cell & B Cell Receptor (BCR) Analysis | • CITE-seq & BCR sequencing<br>• DEG & GSEA<br>• Somatic Hypermutation (SHM) & clonal diversity analysis | • While B cell subset frequencies did not change significantly, Converters showed an induction of age-associated B cell (ABC) transcriptional signatures before RA onset.<br>• B cells from Converters displayed lower clonal diversity than Nonconverters at both baseline and follow-up, suggesting antigen-driven selection. |
| NK & Myeloid Cell Profiling | • Mass Cytometry & CITE-seq<br>• GLMMs for cell frequency associations | • Mature (CD56$^{dim}$CD16$^{+}$CD57$^{+}$) and activated (CD56$^{dim}$CD16$^{+}$CD137$^{+}$) NK cells were expanded in Converters at follow-up.<br>• Myeloid classical monocytes (cM) were expanded and dendritic cells (mDCs) were depleted in Converters at follow-up. |

| | | |
|---|---|---|
| Epigenetic Profiling (Chromatin Accessibility) | • Single-cell ASAP-seq (scASAP-seq)<br>• Differential accessibility analysis | • Converters showed widespread epigenetic differences compared to Nonconverters, with myeloid and NK cells contributing the most changes at baseline and follow-up. |
| Predictive Modeling | • Random Forest, Decision Tree, & Cox Proportional Hazards models<br>• Kaplan-Meier survival analysis | • A model combining baseline cellular features (Tph, *GZMK*$^+$*XCL1*$^+$ CD8$^+$ T cells, and *GZMB*$^+$*CD57*$^+$ CD8$^+$ T cells) and clinical data (anti-CCP3 levels, RF-positivity, and dosage of shared epitope alleles in HLA loci) provided the best prediction for future RA conversion (AUC = 0.763).<br>• Higher baseline frequencies of Tph cells and higher anti-CCP3 titers were significantly associated with a shorter time to RA onset in Converters. |

**Independent analysis of T cell subsets by CITE-seq and mass cytometry data**

In the main text, we present an integrated analysis of T cell populations to maximize statistical power and leverage multi-omic annotations. However, to first characterize the T cell dynamics within each high-parameter platform independently, we performed separate, fine-resolution clustering on datasets from both CITE-seq and mass cytometry. This initial analysis identified key CD4$^+$ and CD8$^+$ T cell subclusters, including peripheral helper T (Tph) cells, *CXCR5*$^+$ CD4$^+$ (Tfh) cells, regulatory T cells (Tregs), and cytotoxic subsets such as *GZMK*$^+$ and *GZMB*$^+$ CD8$^+$ T cells (**Supplementary Figure 2a,d**).

Analysis of the CITE-seq data using generalized linear mixed models (GLMMs) revealed significant alterations in RA Converters before disease onset. At baseline, *GZMK*$^+$*XCL1*$^+$ and *GZMK*$^+$ CD8$^+$ T cells were significantly enriched in Converters compared to Nonconverters (odds ratio (OR) [95% confidence interval (CI)] = 1.44 [1.10-1.88] and 1.31 [1.01-1.88], respectively) (**Supplementary Figure 2b,c**). Within the $CD4^+$ T cell compartment, Tph cells and memory $CD4^+$ T cells did not show enrichment at baseline but expanded significantly by the follow-up time point (OR [95%CI] = 1.57 [1.05-2.36] for Tph cells; OR [95%CI] = 1.31 [1.04-1.65] for memory $CD4^+$ T cells), suggesting a progressive expansion during the transition to clinical RA.

To validate these findings, we assessed T cell associations in our larger mass cytometry cohort. Although not all individual subsets reached statistical significance across both platforms, we observed a consistent pattern of enrichment for Tph cells and activated $CD8^+$ T cells in Converters at both baseline and follow-up (**Supplementary Figure 2e,f**). The general concordance between the two platforms supported the robustness of our findings and provided the rationale for the cross-platform integration detailed in the main text, which allowed us to combine the deep phenotyping of CITE-seq with the larger sample size of the mass cytometry data.

**Integration of CITE-seq and mass cytometry data**

To facilitate cross-platform comparisons of immune profiles between Converters and Nonconverters, we projected the mass cytometry data into the CITE-seq reference space using 33 shared surface proteins enriched for T cell markers (**Methods**). Cluster correspondence was confirmed by match-rate heatmaps (**Supplementary Figure 2g**), which showed strong alignment for major $CD4^+$ and $CD8^+$ T cell subsets, such as Tph, naive and memory $CD4^+$ T cells, and *GZMB*$^+$*CD57*$^+$ $CD8^+$ T cells, supporting the robustness of cross-modality cluster label transfer. While alignment for *GZMK*$^+$ $CD8^+$ T cells and *GZMK*$^+$*XCL1*$^+$ $CD8^+$ T cells were understandably constrained by the lack of GZMK antibody in the mass cytometry panel, the memory $CD8^+$ T cell subset in mass cytometry data showed the highest correspondence with the *GZMK*$^{+}$*XCL1*$^{+}$ CD8$^{+}$ population, and both populations were expanded in Converters at baseline. This integrative approach enabled us to leverage the larger sample size, and thus greater statistical power, of the mass cytometry dataset, while utilizing the detailed cell type annotations provided by CITE-seq, which measures both mRNA and surface proteins, to guide and interpret immune subset classification across both platforms.

**Effect of hydroxychloroquine (HCQ)**

While the StopRA trial concluded that hydroxychloroquine (HCQ) did not prevent the onset of clinical rheumatoid arthritis (RA), our detailed immunological analysis provides insights into its modulatory effects on peripheral immune cell subsets during the preclinical phase. By stratifying our analyses by treatment arm (HCQ vs. placebo), we observed several treatment-associated alterations in T cells, B cells, and NK cells, which may explain the immunomodulatory mechanisms of HCQ noted in established RA and inform the design of future prevention studies.

Our analysis of T cell subsets revealed that the expansion of T peripheral helper (Tph) and memory CD4$^{+}$ T cells, which was significantly associated with RA conversion at the follow-up time point, was only observed in the placebo group. This association was absent in the HCQ-treated group, suggesting that HCQ may attenuate the proliferation or accumulation of these key pathogenic cell types (OR [95%CI] = 1.86 [1.0-3.29] for Tph cells in the placebo group; OR [95%CI] = 1.22 [0.91-1.62] for Tph cells in the HCQ group; OR [95%CI] = 1.42 [1.13-1.79] for memory CD4$^{+}$ T cells in the placebo group; OR [95%CI] = 1.09 [0.84-1.43] for memory CD4$^{+}$ T cells in the HCQ group) (**Supplementary Figure 4a-b**). Furthermore, HCQ treatment was associated with lower levels of cytotoxic *GZMB*$^{+}$*CD57*$^{+}$ CD8$^{+}$ T cells in both Converters and Nonconverters. A significant longitudinal decrease in this population was specifically noted in HCQ-treated Nonconverters (paired Wilcoxon signed-rank test, *p*-value = 0.0096;

**Supplementary Figure 4c**). While the StopRA trial showed that HCQ did not prevent onset of clinical RA, these findings raise the possibility that HCQ may attenuate immune cell alterations associated with disease progression, yet in an insufficient manner to affect the clinical outcome. This could provide a mechanistic explanation for its immunomodulatory effects that have previously been in individuals with established clinical RA[1–3].

We observed a similar modulatory effect on B cell subsets. Among individuals who converted to RA, those treated with HCQ had a significantly higher proportion of naïve B cells (the Wilcoxon rank-sum test, *p*-value = 6.4e-05) and a lower proportion of memory B cells (the Wilcoxon rank-sum test, *p*-value = 2.6e-04) at follow-up compared to those who received the placebo (**Supplementary Figure 7o**). This suggests HCQ may impact B cell differentiation and maturation, potentially restraining the development of memory B cell populations that contribute to autoimmunity.

In the NK cell compartment, the frequency of $CD56^{dim}CD16^{+}$ NK cells was significantly higher in Converters than Nonconverters at follow-up, but this difference was only statistically significant within the placebo group (**Supplementary Figure 9b**). This finding implies that the expansion of this NK cell subset is associated with RA progression in the absence of treatment, and HCQ may mitigate this change.

In summary, while HCQ did not prevent the clinical onset of RA in the StopRA trial, our multi-omic analysis demonstrates that it measurably attenuates pro-inflammatory shifts in T, B, and NK cell compartments that are associated with disease conversion. These findings underscore the importance of considering treatment context when developing and applying biomarkers for risk stratification in preclinical RA.

**TCR and BCR usage analysis**

To investigate TCR usage patterns, we performed comprehensive VDJ gene segment analysis for $CD4^+$ and $CD8^+$ T cells. The proportion of public TCRs, those that are shared across individuals and often respond to common antigens such as Epstein-Barr virus and cytomegalovirus[4], was not significantly different between groups (**Supplementary Figure 6a**). V, D, and J segments showed subtle shifts in usage between Converters and Nonconverters, such as higher *TRBV10-3* in $CD4^+$ T cells in Converters at baseline (**Supplementary Figure 6b-c** for $CD4^+$ T cells, **Supplementary Figure 6f-g** for $CD8^+$ T cells). These differences were visualized in the global view via UMAP projection and their kernel density estimation (KDE) plots incorporating overall VDJ gene usages, where group separation was modest but evident (**Supplementary Figure 6d-e** for $CD4^+$ T cells**, Supplementary Figure 6h-i** for $CD8^+$ T cells), suggesting underlying convergence in TCR features within Converters.

In terms of BCR V(D)J gene usage, no clear difference was observed across groups and time points (**Supplementary Figure 8a-e**), including *IGHV4-34*, an immunoglobulin heavy chain variable gene associated with autoreactive B cell responses[5–7], which may reflect that *IGHV4-34* is associated with anti-CCP antibody status rather than directly distinguishing Converters from Nonconverters.

#### Supplementary Notes

##### Collection and processing of samples

PBMCs were isolated from whole blood collected at each visit. The blood samples were processed using standard density gradient centrifugation protocols at designated biorepository sites (OMRF and University of Colorado). A single aliquot of 10 million PBMCs per sample was allocated for multi-omic profiling, including CITE-seq, mass cytometry (CyTOF), scASAP-seq, and T/B cell receptor sequencing. All PBMC, plasma, DNA, and RNA samples were processed and cryopreserved at −80°C or in liquid nitrogen vapor phase at the Oklahoma Medical Research Foundation (OMRF), the central biorepository for the StopRA trial. Additional serum and urine samples were stored at the University of Colorado. Samples were logged and tracked using standardized inventory systems in collaboration with the Statistical and Clinical Coordinating Center for Autoimmune Diseases Clinical Trials (SACCC-ADCT). Strict quality control measures were implemented to ensure sample viability for downstream multi-omic and proteomic analyses.

##### Preprocessing of single-cell multi-omic assays

We provided a list of antibodies for each assay as **Supplementary Table 2**.

*Sample processing for CITE-seq/scTCR-seq/scBCR-seq/scASAP-seq*

Single-cell transcriptome and immune receptor profiling were performed using the 10x Genomics Single Cell V(D)J and 5′ Gene Expression solutions. For each sample, PBMCs were stained with a TotalSeq-C panel of 137 barcoded antibodies (BioLegend), followed by microfluidic encapsulation on the Chromium X platform (10x Genomics). Parallel droplet generation enabled simultaneous capture of 5′ scRNA-seq, surface protein expression (CITE-seq), and paired V(D)J sequences for TCR and BCR. Separate libraries were prepared for gene expression, ADT (antibody-derived tags), and immune repertoire libraries (TCR/BCR) following the manufacturer's protocols. Sequencing was performed on an Illumina NovaSeq platform to a target depth of ~30,000–50,000 reads per cell for gene expression and ~5,000 reads per cell for ADTs.

*Preprocessing and quality control steps for CITE-seq*

Raw BCL files were demultiplexed using the *mkfastq* tool within CellRanger software (10X Genomics, version 8.0.0) with default parameters to generate FASTQ files. Subsequent to this, the samples were aligned to the human reference genome (GRCh38), and gene counts were quantified using the *count* tool within CellRanger software. To correct for ambient RNA contamination and improve signal-to-noise in CITE-seq data, raw gene expression matrices were processed using *CellBender remove-background* (v0.3.0). The --fpr 0.1 parameter was specified based on recommendations from the original CellBender publication for CITE-seq datasets to balance sensitivity and specificity[8]. Initial quality control excluded cells with low complexity (fewer than 500 detected genes) or high mitochondrial RNA content (≥20%). Samples with fewer than 300 high-quality cells were also removed. Doublet detection was performed using the scDblFinder package (v1.16.0)[9], and predicted doublets were excluded from downstream analysis. Following QC, gene expression matrices were normalized using log-transformation, and highly variable genes were identified after excluding mitochondrial, ribosomal, and cell cycle–related genes. Dimensionality reduction was achieved via canonical correlation analysis (CCA) using the PMA R package[10] followed by Harmony-based batch correction[11] (v1.2.0) across both batch and sample ID metadata. LISI (Local Inverse Simpson's Index) scores were calculated to evaluate batch integration quality across CCA and Harmony spaces. Final clustering and dimensionality reduction embeddings were stored as Seurat objects and used for downstream lineage-specific analyses. Uniform manifold approximation and projection (UMAP) was applied to the top 20 Harmony-corrected canonical variates for visualization. To classify major immune lineages from peripheral blood mononuclear cells (PBMCs), we utilized normalized antibody-derived tag (ADT) data from CITE-seq. Protein expression values were normalized using the cellCLR method, and lineage-specific thresholds were applied to distinguish T cells, B cells, NK cells, and myeloid cells based on canonical surface protein markers; CD3 (T cells), CD19 (B cells), CD56 (NK cells), and CLEC12A (myeloid cells). After lineage classification, unsupervised clustering was performed within each lineage subset using the Seurat package (v5.0.3)[12]. Louvain clustering was conducted using the shared nearest neighbor (SNN) graph, with the following lineage-specific resolution parameters: 1 for T cells, 0.20 for myeloid cells, 0.40 for B cells, and 1 for NK cells. The clustering resolution for each lineage was selected empirically to balance granularity and interpretability, ensuring biologically meaningful subpopulation separation. We assigned biological cluster annotation names for each louvain cluster based on protein and mRNA expression patterns.

*Preprocessing and quality control steps for scTCR-seq and scBCR-seq*

For the immune repertoire analysis, FASTQ files using CellRanger software (10X Genomics, version 8.0.0) with default parameters were processed using the *vdj* tool within CellRanger to assemble sequences and identify clonotypes for both T-cell receptors (TCR) and B-cell receptors (BCR) across the samples. Each sample was analyzed for TCR sequences using the filtered_contig_annotations.csv file generated by CellRanger. First we removed cells with multiple heavy chains. The identified TCR and BCR sequences, along with their corresponding cell barcodes, were then aligned with cell barcodes from the CITE-seq library of the same sample. For downstream analysis, we included TCR and BCR with productive sequences. Clonal calls were performed by the combination of the nucleotide and gene sequence using “strict” in each function of the scRepertoire package.

*Preprocessing and quality control steps for scASAP-seq*

PBMCs were profiled using single-cell ASAP-seq, which captures chromatin accessibility and surface protein expression (ADT) from the same cells. Initial preprocessing involved merging Seurat objects from individual samples and performing quality control (QC) filtering. Cells were retained if they met all of the following thresholds: total fragments (nCount_ATAC) between 1,000 and 100,000, TSS enrichment score > 2, nucleosome signal < 4, fraction of reads in blacklist regions < 0.0001, and percent of reads in peaks > 10. For dimensionality reduction, ATAC-seq data were normalized using term frequency-inverse document frequency (TF-IDF) followed by latent semantic indexing (LSI), while ADT data were normalized using centered log-ratio (CLR) transformation and reduced using PCA. To integrate chromatin accessibility and protein expression modalities, we applied CCA. UMAP was applied to the top 20 Harmony-corrected principal components for visualization. Louvain clustering was then applied to the batch-corrected CCA embeddings at resolution 0.20. Cell types were assigned to clusters based on both ADT marker expressions (CD3, CD4, CD8, CD19, CD56, CD14, and CD11b). Peak calling was performed by sub-clusters in each board cell type using macs2 (v 2.2.9.1).

*Sample processing for mass cytometry*

After Fc receptors were blocked using Human TruStain FcX (BioLegend) for 10 minutes at room temperature, cells were then stained with the T Cell Expansion Panel 3 and Maxpar Direct

Immune Profiling Assay (MDIPA) antibodies (Standard BioTools). Antibodies in the T Cell Expansion Panel were titrated beforehand using a 10-tube titration method to minimize signal spillover, and final dilutions were optimized to ensure consistent marker resolution across individuals ￼. Cells were fixed using freshly prepared 1.6% paraformaldehyde and permeabilized with Maxpar Perm-S buffer. Intracellular staining was then performed using the Basic Activation Panel, which includes antibodies targeting cytokines and cytotoxic markers such as Granzyme B, Perforin, IFN-γ, and TNF-α. Following two washes with cell staining buffer, cells were stained overnight at 4°C with 125 nM Cell-ID Intercalator-Ir for DNA content normalization. After intercalator staining, cells were washed, counted again, and resuspended in Cell Acquisition Solution containing Benzonase. All samples were acquired using the CyTOF XT instrument, targeting approximately 200,000 events per sample. Instrument settings were standardized using established templates and dilution parameters to maintain consistency across batches.

*Preprocessing and quality control steps for mass cytometry*

After acquisition, FCS files were exported and preprocessed using standard workflows. Quality control involved the removal of debris and doublets based on event length and DNA intercalator intensity. Events with abnormal Ir191/Ir193 signal intensities were excluded. Bead-based normalization and compensation for signal spillover were performed as needed. Clustering and dimensionality reduction were conducted on arcsinh-transformed data using canonical markers to identify major immune cell populations. Markers showing high spillover or poor resolution were manually inspected, and cells with ambiguous marker expression profiles were excluded from downstream analyses. We applied lineage-based gating to classify cells into major immune compartments: CD3 (T cells), CD19 (B cells), CD56 (NK cells), and CD14 (myeloid cells). Principal component analysis (PCA) was applied to the normalized and scaled expression data, and batch correction was performed using Harmony, adjusting for batch and sample index.

UMAP was computed from the top 20 Harmony components. To identify cell clusters, a SNN graph was constructed using the top Harmony embeddings, followed by Louvain modularity optimization at optimized resolution parameters: 0.60 for T cells, 0.20 for myeloid cells, 0.20 for B cells, and 0.20 for NK cells. The clustering resolution for each lineage was selected empirically to balance granularity and interpretability, ensuring biologically meaningful subpopulation separation. We assigned biological cluster annotation names for each louvain cluster based on protein expression patterns.

**Geneset of signatures**

For Tph-signature[13] , *CXCL13, RNF19A, DUSP4, PDCD1, TOX2, MAF, CTLA4, TSHZ2, TIGIT, SNX9, PTPN13, NR3C1, ITM2A, FKBP5, TP53INP1, ARID5B, TOX, COTL1, AHI1*, and *ZNRF1*, were included.

For *CD96*$^{high}$ CD4$^{+}$ T cell signature[14], *AHR, AIRE, ALOX5AP, ANKRD28, ANKRD50, APOE, BHLHE40, CAPG, CCR6, CD63, CD96, CFH, CLU, CMTM6, CNTNAP1, CRIP2, CTSH, CYB561, DENND2C, EEF1DP3, F3, FA2H, FAM189A2, FXYD7, GAB3, GLIPR1, GLUL, GPR65, IGF2BP3, IL22, IL4I1, ITGAE, KIT, LGALS3, LZTS1, MGLL, NR1D1, NT5E, OR13A1, PGAP4, PLEKHG3, PLP2, PPP3CA, PRNP, RHOQP1, RIMS2, RORC, RPS6KA3, S100A6, SASH1, SEPTIN11, SERHL, SLC14A2, SOS1, SRPX, SYTL2, TIMP1, TMPRSS11E, TNFRSF11A, TNFSF13B, TRAPPC3L, TREML1, USP43, VHLL, VIM, WDR86-AS1, ZDHHC11, ZNF705EP,* and *ZNRF3-AS1* were included.

For Treg signature[13], *FOXP3, TIGIT, IKZF2, DUSP4, CTLA4, ARID5B, STK17B, PELI1, F5, RGS1, PKM, CD4, ENTPD1, YWHAB, IL32, IKZF4, BIRC3, AC013394.2, GBP2,* and *SNX9* were included.

For Th17 signature[15], *AATK, ABCA1, ABCB1, ADAM10, AHCY, ALG1, AMMECR1, ANK1, ANO6, AP3S2, ATF5, BCCIP, C16orf74, C2CD4B, C4orf46, CASR, CCDC6, CEBPG, CEP97, CERK, CFL2, CMTM6, COMMD3, CR1, CTLA4, DAB1, DNAJC10, DST, DUSP16, EXT1, FAM126A, FAM63B, FBL, FRMD4A, FURIN, FUT10, GDF7, GMDS, GRAMD3, GTF3C4, HADHA, HERC6, HRH4, IKZF3, IL12RB1, IL1R2, INPP5F, JMY, KLHL42, KLHL5, LDLRAD4, LSM10, LTA4H, LTB, MAN1A1, MAP3K4, MGAT4A, MIEN1, MRPL34, NPAS2, PALB2, PANK4, PDDC1, PDE4D, PHF21A, PLXNC1, PPA1, PPIF, PRKRIR, PSD3, PSMG3, PTPN4, RAD54B, RANBP9, RPLP0, SBK1, SEMA6C, SESN1, SKA3, SLAMF1, SLC35F2, SLC35G1, SOCS2, SRD5A3, SYNGAP1, TBC1D4, TLR2, TMED8, TMEM156, TMEM60, TP53INP1, TRAF3IP2, UBAP1, UFC1, UROS, USP10, YEATS4, YWHAH,* and *ZC3H12D* were included.

For Th1 signature[15], *ABHD17A, ABI3, ACRC, ACSL3, ADAMTS10, AGPAT5, AKNA, APMAP, APPBP2, ARAP2, ARHGAP26, ARPC5L, ASAH2B, ATP10A, BACH2, BBC3, BCR, BTBD11, BTN3A1, C9orf91, CARHSP1, CBLB, CCDC64, CCDC78, CCNDBP1, CDCA7L, CDK5R1, CLHC1, CLIC6, CLN8, CMC1, CMPK2, CNN3, COL6A2, CRTAM, CTSW, CXCR3, DFNB31, DIP2A, DKK3, DOCK3, DRAM1, DTHD1, E2F3, ENC1, ENG, ENTPD6, EOMES, EPB41L5, EXPH5, F2R, FCRL6, FGFBP2, FLT4, FOXJ1, FYN, GBP4, GBP5, GFOD1, GGT7, GNB3, GSE1, GZMK, GZMM, HAVCR1, HBEGF, HIC1, IFI27, IFNG, IGF1R, IGFBP3, IGFBP4, IL12RB2, INPP5A, IRF8, IVD, JAK2, KIAA1522, KIAA1671, L3MBTL4, LDLRAP1, LGR6, LITAF, LMO7, LPCAT1, LUZP1, LYAR, MAN2A1, MANBA, MAPRE3, MATK, MCTP2, ME3, MPP1, MZB1, NCALD, P3H3, PACSIN1, PDE9A, PDSS1, PDZD8, PHLPP1, PMAIP1, PMEPA1, PRF1, PRMT2, PSME4, PTK2B, PTPRE, PTPRJ, PWP2, QSOX2, RAB24, RASGEF1B, RASGRF2, RHBDF2, ROBO3, ROR2, RRAGD, RSAD2, RTP5, RUFY4, RUSC2, S100PBP, S1PR5, SAMD3, SAV1, SCD, SH2B3, SH3RF3, SIPA1, SIRT3, SLA2, SLAMF7, SLC4A4, SMAD3, SMAD7, SNTB1, SOS2, SPATA20, SRPK3, SYNM, TBKBP1, TBX21, TMEM109, TMEM62, TNS4, TRAPPC10, TRPS1, TSPAN33, TTC23, TULP4, UPP1, XCL1,*

*YARS, YES1, ZBTB49, ZC4H2, ZCCHC18, ZIK1, ZNF135, ZNF541, ZNF618, ZNF629,* and *ZNF711* were included.

For the ABC-gene signature[16], *CD40, CD86, FAS, FCRL4, FCRL5, FCRLB, FGL2, CXCL10, CXCR3, NKG7, ITGAM, ITGAX, ITGB2, TBX21, ZBTB32, FCER2, CR2,* and *ZEB2* were included.

#### Supplementary References


1. Takei, H. *et al.* Clinical and immunological effects of hydroxychloroquine in patients with active rheumatoid arthritis despite antirheumatic treatment. *Mod Rheumatol* **34**, 50–59 (2023).
2. Rempenault, C. *et al.* Clinical and Structural Efficacy of Hydroxychloroquine in Rheumatoid Arthritis: A Systematic Review. *Arthritis Care Res (Hoboken)* **72**, 36–40 (2020).
3. Alam, M. K. *et al.* Comparative study on methotrexate and hydroxychloroquine in the treatment of rheumatoid arthritis. *Mymensingh Med J* **21**, 391–398 (2012).
4. Li, H., Ye, C., Ji, G. & Han, J. Determinants of public T cell responses. *Cell Research* **22**, 33–42 (2012).
5. Pugh-Bernard, A. E. *et al.* Regulation of inherently autoreactive VH4-34 B cells in the maintenance of human B cell tolerance. *J Clin Invest* **108**, 1061–1070 (2001).
6. Tipton, C. M. *et al.* Diversity, cellular origin and autoreactivity of antibody-secreting cell population expansions in acute systemic lupus erythematosus. *Nat Immunol* **16**, 755–765 (2015).
7. Cowan, G. J. M. *et al.* In Human Autoimmunity, a Substantial Component of the B Cell Repertoire Consists of Polyclonal, Barely Mutated IgG B Cells. *Front Immunol* **11**, 395

(2020).

8. Fleming, S. J. *et al.* Unsupervised removal of systematic background noise from droplet-based single-cell experiments using CellBender. *Nature Methods* **20**, 1323–1335 (2023).
9. Germain, P.-L., Lun, A., Garcia Meixide, C., Macnair, W. & Robinson, M. D. Doublet identification in single-cell sequencing data using. *F1000Res* **10**, 979 (2021).
10. GitHub - bnaras/PMA. *GitHub* https://github.com/bnaras/PMA.
11. Korsunsky, I. *et al.* Fast, sensitive and accurate integration of single-cell data with Harmony. *Nat Methods* **16**, 1289–1296 (2019).
12. Hao, Y. *et al.* Dictionary learning for integrative, multimodal and scalable single-cell analysis. *Nature Biotechnology* **42**, 293–304 (2023).
13. Zhang, F. *et al.* Defining inflammatory cell states in rheumatoid arthritis joint synovial tissues by integrating single-cell transcriptomics and mass cytometry. *Nature Immunology* **20**, 928–942 (2019).
14. Law, C. *et al.* Interferon subverts an AHR–JUN axis to promote CXCL13+ T cells in lupus. *Nature* **631**, 857–866 (2024).
15. Höllbacher, B. *et al.* Transcriptomic Profiling of Human Effector and Regulatory T Cell Subsets Identifies Predictive Population Signatures. *Immunohorizons* **4**, 585–596 (2020).
16. Dai, D. *et al.* The transcription factor ZEB2 drives the formation of age-associated B cells. *Science* **383**, 413–421 (2024).

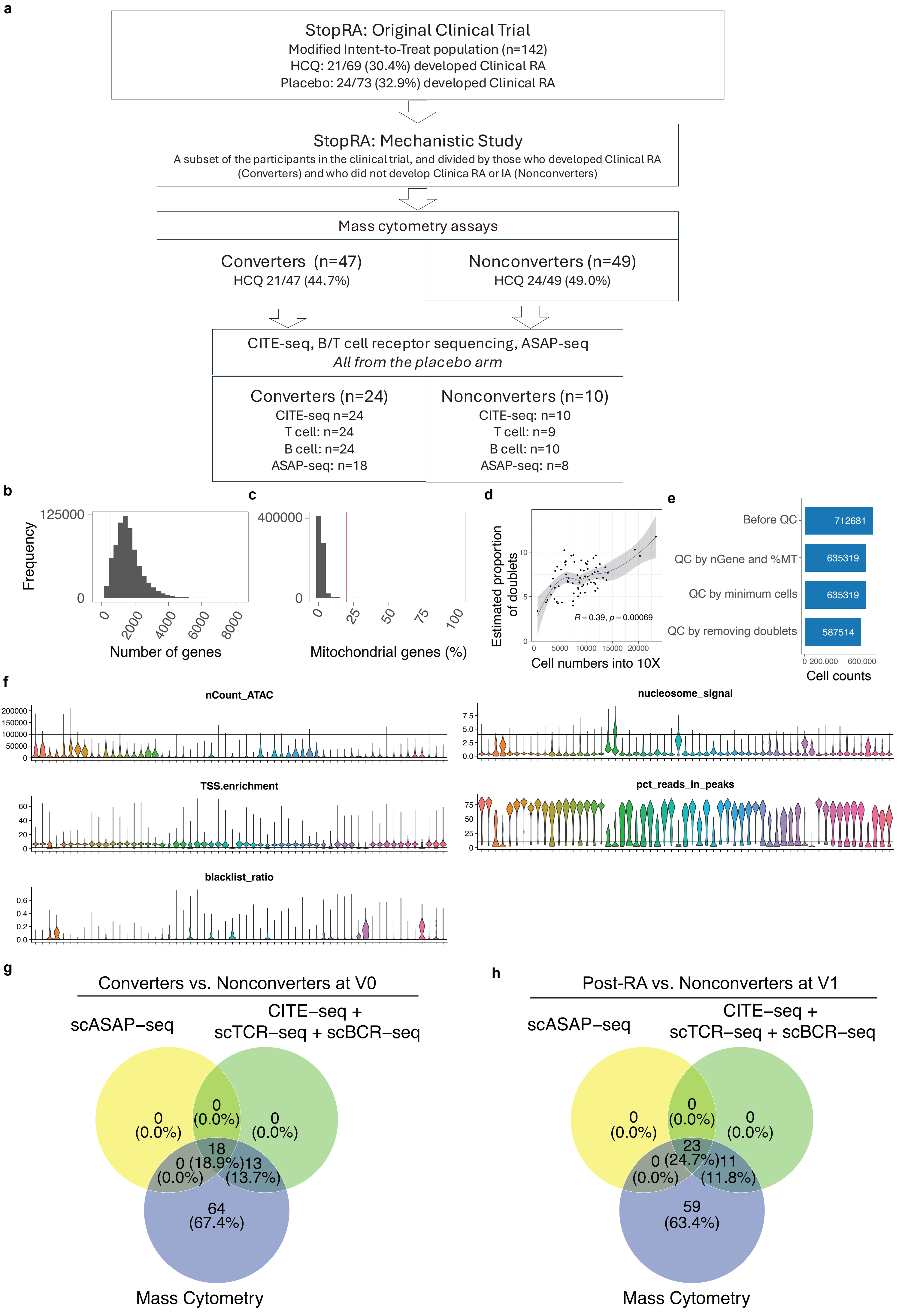
a
StopRA: Original Clinical Trial
Modified Intent-to-Treat population (n=142)
HCQ: 21/69 (30.4%) developed Clinical RA
Placebo: 24/73 (32.9%) developed Clinical RA
StopRA: Mechanistic Study
A subset of the participants in the clinical trial, and divided by those who developed Clinical RA (Converters) and who did not develop Clinica RA or IA (Nonconverters)
Mass cytometry assays
Converters (n=47)
HCQ 21/47 (44.7%)
Nonconverters (n=49)
HCQ 24/49 (49.0%)
CITE-seq, B/T cell receptor sequencing, ASAP-seq
All from the placebo arm
Converters (n=24)
CITE-seq n=24
T cell: n=24
B cell: n=24
ASAP-seq: n=18
Nonconverters (n=10)
CITE-seq: n=10
T cell: n=9
B cell: n=10
ASAP-seq: n=8
b
Frequency
Number of genes
c
Mitochondrial genes (%)
d
Estimated proportion of doublets
Cell numbers into 10X
R = 0.39, p = 0.00069
e
Before QC
QC by nGene and %MT
QC by minimum cells
QC by removing doublets
712681
635319
635319
587514
Cell counts
f
nCount_ATAC
nucleosome_signal
TSS.enrichment
pct_reads_in_peaks
blacklist_ratio
g
Converters vs. Nonconverters at V0
scASAP−seq
CITE−seq + scTCR−seq + scBCR−seq
Mass Cytometry
0 (0.0%)
0 (0.0%)
0 (0.0%)
18 (18.9%)
0 (0.0%)
13 (13.7%)
64 (67.4%)
h
Post-RA vs. Nonconverters at V1
scASAP−seq
CITE−seq + scTCR−seq + scBCR−seq
Mass Cytometry
0 (0.0%)
0 (0.0%)
0 (0.0%)
23 (24.7%)
0 (0.0%)
11 (11.8%)
59 (63.4%)

**Supplementary Figure 1: Quality control (QC) across multi-omics modalities. a**, Study design and sample selection. Flow chart illustrating the sub-sampling of the StopRA clinical trial for downstream mechanistic assays. Participants were classified as Converters (developed clinical RA) or Nonconverters. Mass cytometry included participants from both treatment arms, while high-dimensional single-cell sequencing was performed exclusively on the placebo arm. n represents the number of donors with available data at the baseline time point (V0). **b**, Distribution of the number of detected genes per cell in CITE-seq data prior to QC, with a threshold line used for filtering cells with low detected genes. **c**, Distribution of the percentage of mitochondrial genes per cell in CITE-seq data, with a threshold line used for filtering cells with high mitochondrial content. **d**, Relationship between the number of cells in CITE-seq data loaded into the 10x Chromium platform and the estimated proportion of doublets. **e**, Stepwise filtering of cells in CITE-seq data through QC stages: by number of genes and mitochondrial gene percentage, minimum cell counts, and removal of doublets. **f**, Violin plots showing the distribution of selected QC metrics across samples of scASAP-seq data. (**g-h**) Venn diagrams showing overlap of samples available for mass cytometry, CITE-seq with paired scTCR/BCR-seq, and scASAP-seq after QC for baseline (V0; panel **g**) and post-onset/follow-up (V1; panel **h**) timepoints in Converters vs. Nonconverters. Numbers in parentheses indicate the proportion of samples contributed by each modality.

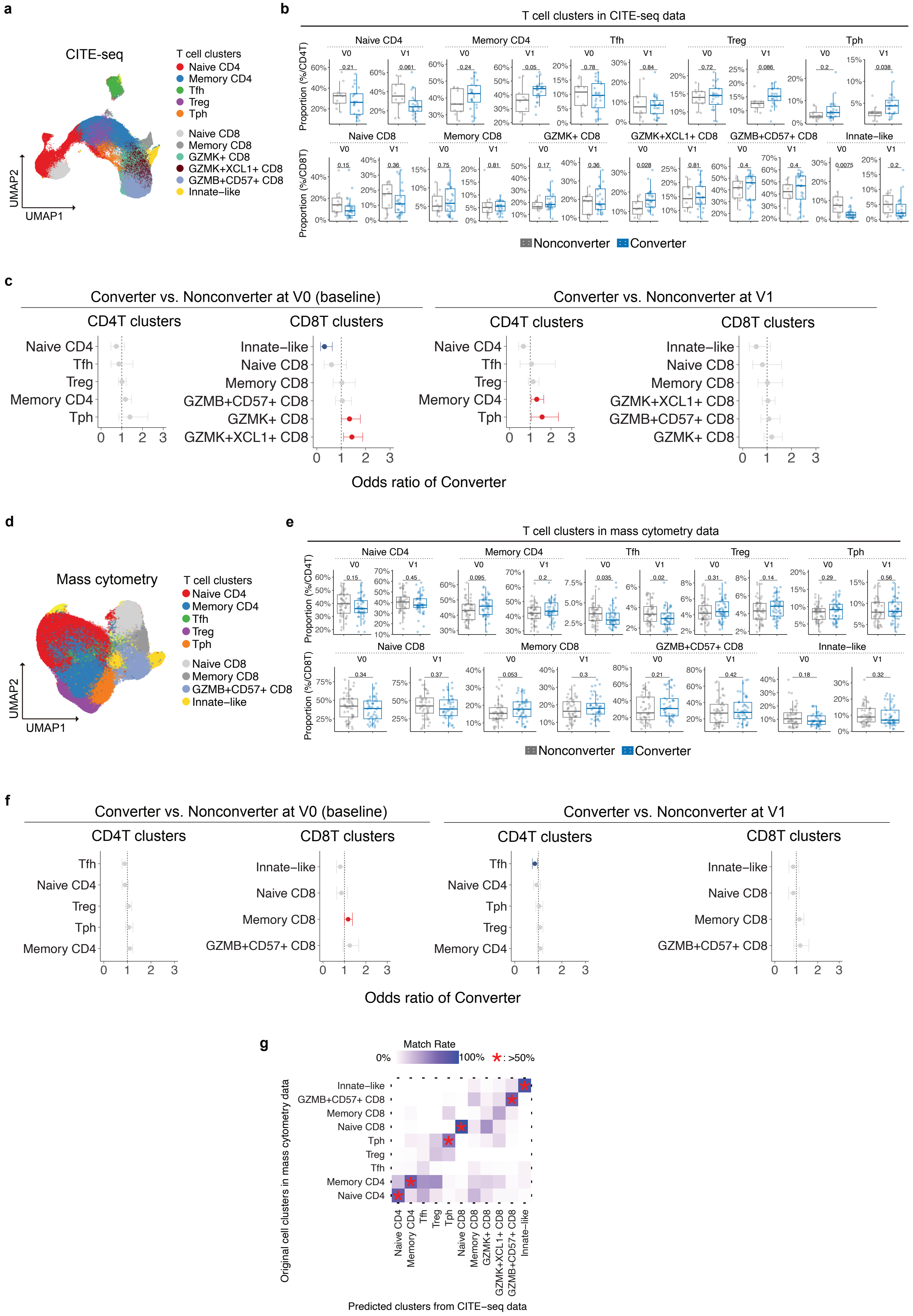

a
CITE-seq
T cell clusters
Naive CD4
Memory CD4
Tfh
Treg
Tph
Naive CD8
Memory CD8
GZMK+ CD8
GZMK+XCL1+ CD8
GZMB+CD57+ CD8
Innate−like
UMAP2
UMAP1
b
T cell clusters in CITE-seq data
Proportion (%/CD4T)
Proportion (%/CD8T)
Naive CD4
Memory CD4
Tfh
Treg
Tph
Naive CD8
Memory CD8
GZMK+ CD8
GZMK+XCL1+ CD8
GZMB+CD57+ CD8
Innate−like
V0
V1
Nonconverter
Converter
c
Converter vs. Nonconverter at V0 (baseline)
Converter vs. Nonconverter at V1
CD4T clusters
CD8T clusters
Odds ratio of Converter
d
Mass cytometry
T cell clusters
Naive CD4
Memory CD4
Tfh
Treg
Tph
Naive CD8
Memory CD8
GZMB+CD57+ CD8
Innate−like
UMAP2
UMAP1
e
T cell clusters in mass cytometry data
Proportion (%/CD4T)
Proportion (%/CD8T)
Nonconverter
Converter
f
Converter vs. Nonconverter at V0 (baseline)
Converter vs. Nonconverter at V1
CD4T clusters
CD8T clusters
Odds ratio of Converter
g
Match Rate
0%
100%
*: >50%
Original cell clusters in mass cytometry data
Predicted clusters from CITE−seq data

**Supplementary Figure 2: Independent analysis of T cell subsets by CITE-seq and mass cytometry data. a**, Fine-scale cell-types in T cells of CITE-seq dataset on UMAP. **b**, Boxplots showing the frequencies of CITE-seq-defined T cell clusters in Converters and Nonconverters at baseline (V0) and post-onset/follow-up (V1). P-values in boxplots are shown above each comparison and were calculated using Wilcoxon rank-sum tests. **c**, Generalized linear mixed model (GLMM) analysis using CITE-seq T cells comparing RA Converters and Nonconverters at each visit point, at baseline (V0, left) and post-onset/follow-up (V1). The model accounted for age, sex, batch and individuals as covariates. The odds ratio and 95% confidence intervals (CIs) are shown separately for $CD4^{+}$ T and $CD8^{+}$ T cell clusters. CIs entirely above 1 are shown in red, indicating a positive association with RA conversion, while those entirely below 1 are shown in blue, indicating a negative association. **d**, Fine-scale cell-types in T cells of mass cytometry dataset on UMAP. **e**, Boxplots showing the frequencies of mass cytometry–defined T cell clusters in Converters and Nonconverters at V0 and V1. P-values in boxplots are shown above each comparison and were calculated using Wilcoxon rank-sum tests. **f**, GLMM analysis using mass cytometry T cells comparing RA Converters and Nonconverters at each visit point, at baseline (V0, left) and post-onset/follow-up (V1). The model accounted for age, sex, batch and individuals as covariates. **g**, Heatmap showing the match rate (%) between original T cell clusters identified by mass cytometry (rows) and predicted clusters derived from CITE-seq data (columns) in mass cytometry data. Color intensity corresponds to the proportion of cells within each cluster that were mapped to each corresponding CITE-seq cluster. Asterisks (*) indicate cluster pairs with a match rate greater than 50%.

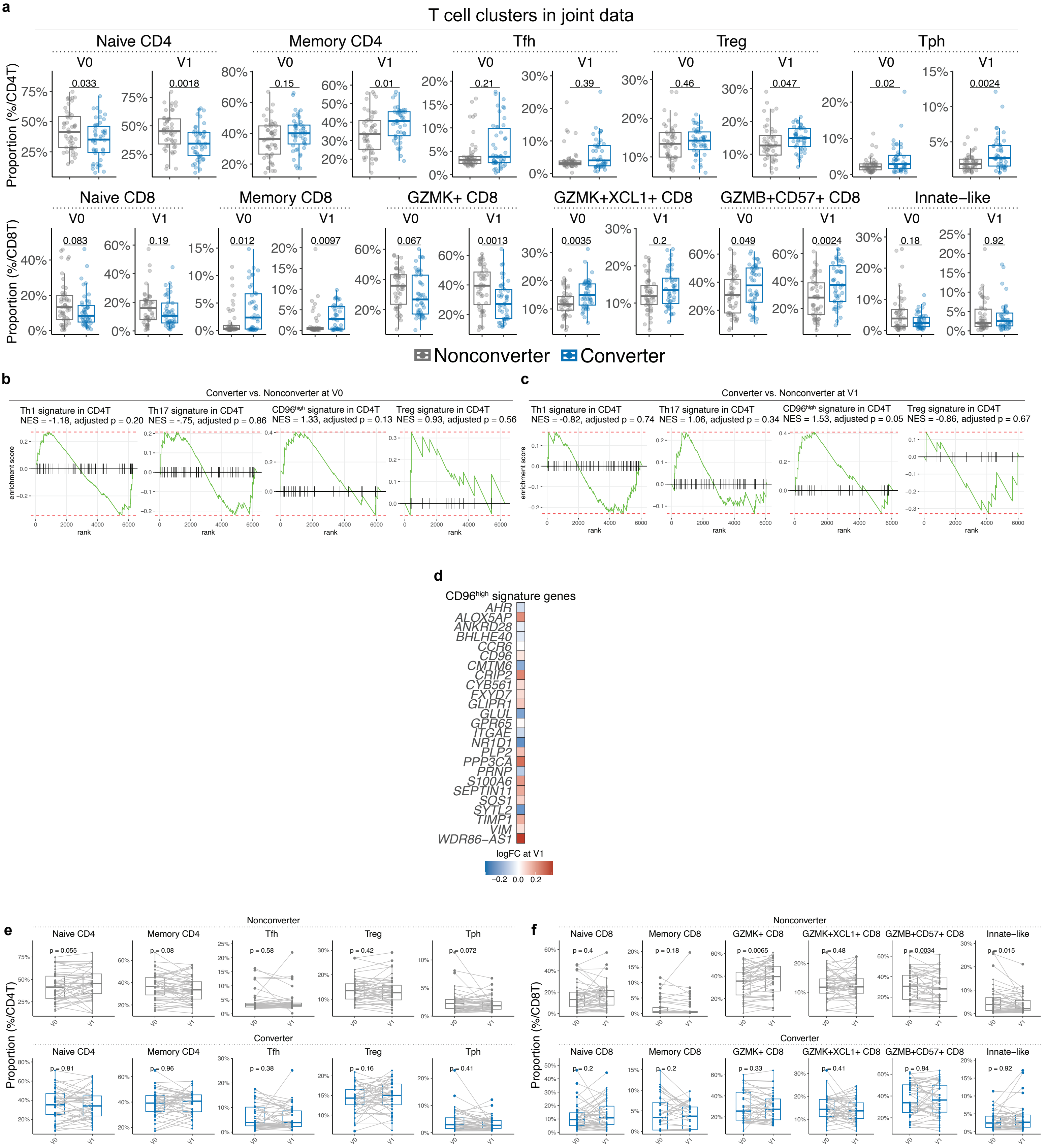
a
T cell clusters in joint data
Naive CD4
Memory CD4
Tfh
Treg
Tph
V0
V1
Proportion (%/CD4T)
0.033
0.0018
0.15
0.01
0.21
0.39
0.46
0.047
0.02
0.0024
Naive CD8
Memory CD8
GZMK+ CD8
GZMK+XCL1+ CD8
GZMB+CD57+ CD8
Innate−like
Proportion (%/CD8T)
0.083
0.19
0.012
0.0097
0.067
0.0013
0.0035
0.2
0.049
0.0024
0.18
0.92
Nonconverter
Converter
b
Converter vs. Nonconverter at V0
Th1 signature in CD4T
NES = -1.18, adjusted p = 0.20
Th17 signature in CD4T
NES = -.75, adjusted p = 0.86
CD96high signature in CD4T
NES = 1.33, adjusted p = 0.13
Treg signature in CD4T
NES = 0.93, adjusted p = 0.56
enrichment score
rank
c
Converter vs. Nonconverter at V1
Th1 signature in CD4T
NES = -0.82, adjusted p = 0.74
Th17 signature in CD4T
NES = 1.06, adjusted p = 0.34
CD96high signature in CD4T
NES = 1.53, adjusted p = 0.05
Treg signature in CD4T
NES = -0.86, adjusted p = 0.67
enrichment score
rank
d
CD96high signature genes
AHR
ALOX5AP
ANKRD28
BHLHE40
CCR6
CD96
CMTM6
CRIP2
CYB561
FXYD7
GLIPR1
GLUL
GPR65
ITGAE
NR1D1
PLP2
PPP3CA
PRNP
S100A6
SEPTIN11
SOS1
SYTL2
TIMP1
VIM
WDR86−AS1
logFC at V1
−0.2 0.0 0.2
e
Nonconverter
Naive CD4
Memory CD4
Tfh
Treg
Tph
p = 0.055
p = 0.08
p = 0.58
p = 0.42
p = 0.072
Converter
p = 0.81
p = 0.96
p = 0.38
p = 0.16
p = 0.41
Proportion (%/CD4T)
f
Nonconverter
Naive CD8
Memory CD8
GZMK+ CD8
GZMK+XCL1+ CD8
GZMB+CD57+ CD8
Innate−like
p = 0.4
p = 0.18
p = 0.0065
p = 0.48
p = 0.0034
p = 0.015
Converter
p = 0.2
p = 0.2
p = 0.33
p = 0.41
p = 0.84
p = 0.92
Proportion (%/CD8T)

**Supplementary Figure 3: Joint analysis of T cell clusters using harmonized data from both CITE-seq and mass data. a,** Boxplots of subset proportions in Converters and Nonconverters across V0 and V1. P-values in boxplots are shown above each comparison and were calculated using Wilcoxon rank-sum tests. **b-c**, Normalized enrichment score (NES) by Gene Set Enrichment Analysis (GSEA) showing the enrichment of the Th1, Th17, $CD96^{high}$, and Treg-signature in Converter at baseline (**b**) and follow-up time point (**c**). **d**, Heatmap showing expression of representative *CD96*$^{high}$ $CD4^+$ T cell-signature genes comparing RA Converters and Nonconverters at follow-up. **e-f**, Longitudinal comparison of $CD4^+$ T cell subsets (**e**) and $CD8^+$ T cell subsets (**f**) frequencies between baseline and follow-up timepoints, stratified by RA conversion status. P-values from paired Wilcoxon signed-rank tests are indicated for each group.

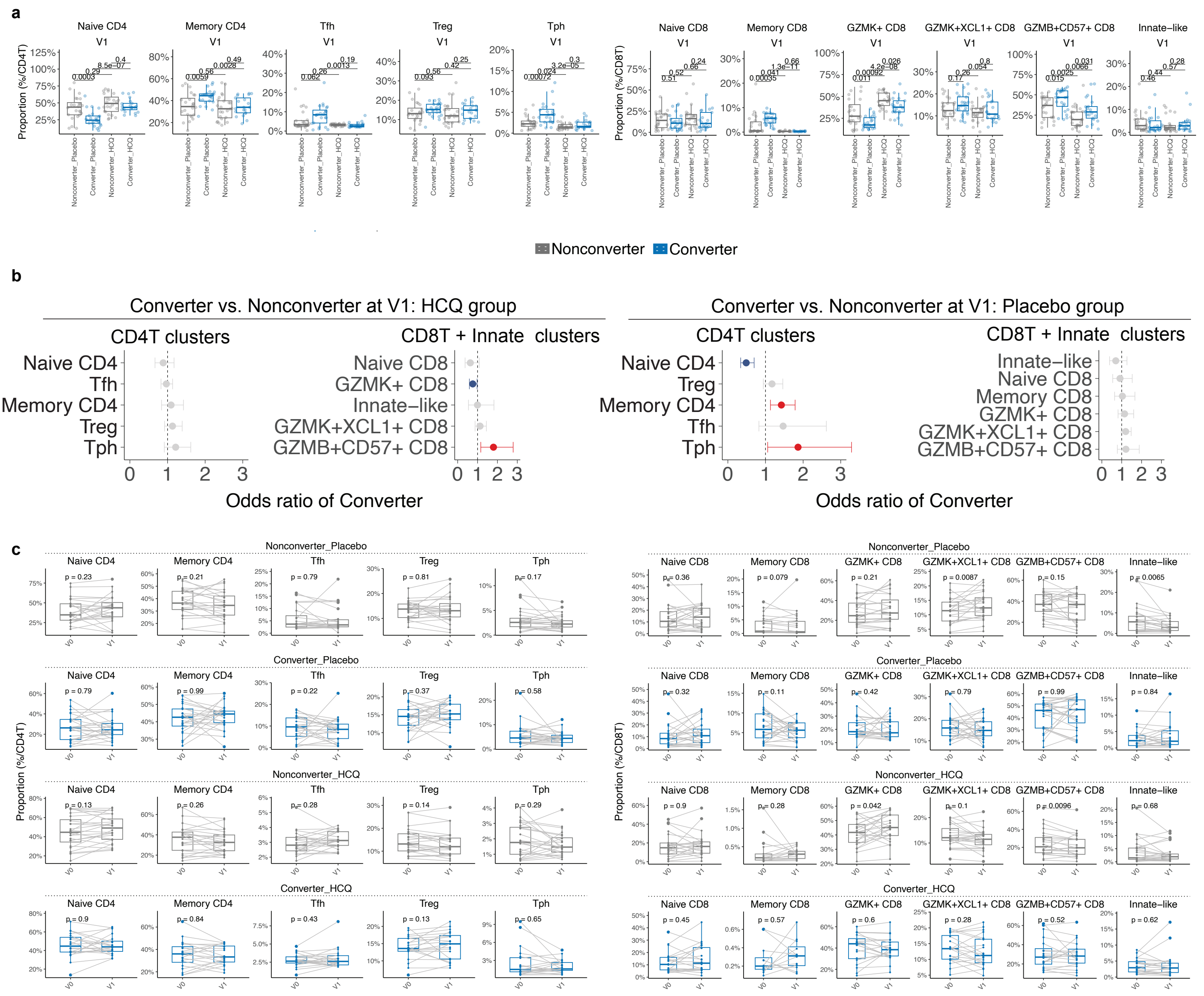
a
Naive CD4
Memory CD4
Tfh
Treg
Tph
V1
Proportion (%/CD4T)
Naive CD8
Memory CD8
GZMK+ CD8
GZMK+XCL1+ CD8
GZMB+CD57+ CD8
Innate−like
Proportion (%/CD8T)
Nonconverter_Placebo
Converter_Placebo
Nonconverter_HCQ
Converter_HCQ
Nonconverter
Converter
b
Converter vs. Nonconverter at V1: HCQ group
CD4T clusters
CD8T + Innate clusters
Converter vs. Nonconverter at V1: Placebo group
Odds ratio of Converter
c
Nonconverter_Placebo
Converter_Placebo
Nonconverter_HCQ
Converter_HCQ
V0
V1

**Supplementary Figure 4: Impact of HCQ treatment on T cell subset frequencies in Converters and Nonconverters. a**, Cross-sectional comparison of T cell subset frequencies at follow-up (V1) between Converters and Nonconverters stratified by treatment arm (Placebo or HCQ). Boxplots display the proportions of $CD4^+$ and $CD8^+$ T cell subsets within total $CD4^+$ or $CD8^+$ T cells, respectively. Gray indicates Nonconverters, and blue indicates Converters. P-values were calculated using Wilcoxon rank-sum tests between groups. **b,** Generalized linear mixed model (GLMM) analysis using CITE-seq T cells comparing RA Converters and Nonconverters at follow-up point (V1) by treatment arms, HCQ group (left) and Placebo group (right). The model accounted for age, sex, batch and individuals as covariates. The odds ratio and 95% confidence intervals (CIs) are shown separately for $CD4^+$ T and $CD8^+$ T cell clusters. CIs entirely above 1 are shown in red, indicating a positive association with RA conversion, while those entirely below 1 are shown in blue, indicating a negative association. **c**, Longitudinal comparison of T cell subset frequencies between baseline (V0) and follow-up (V1) within each treatment and outcome group. Paired boxplots with connecting lines represent intra-individual changes. P-values from paired Wilcoxon signed-rank tests are shown for each comparison.

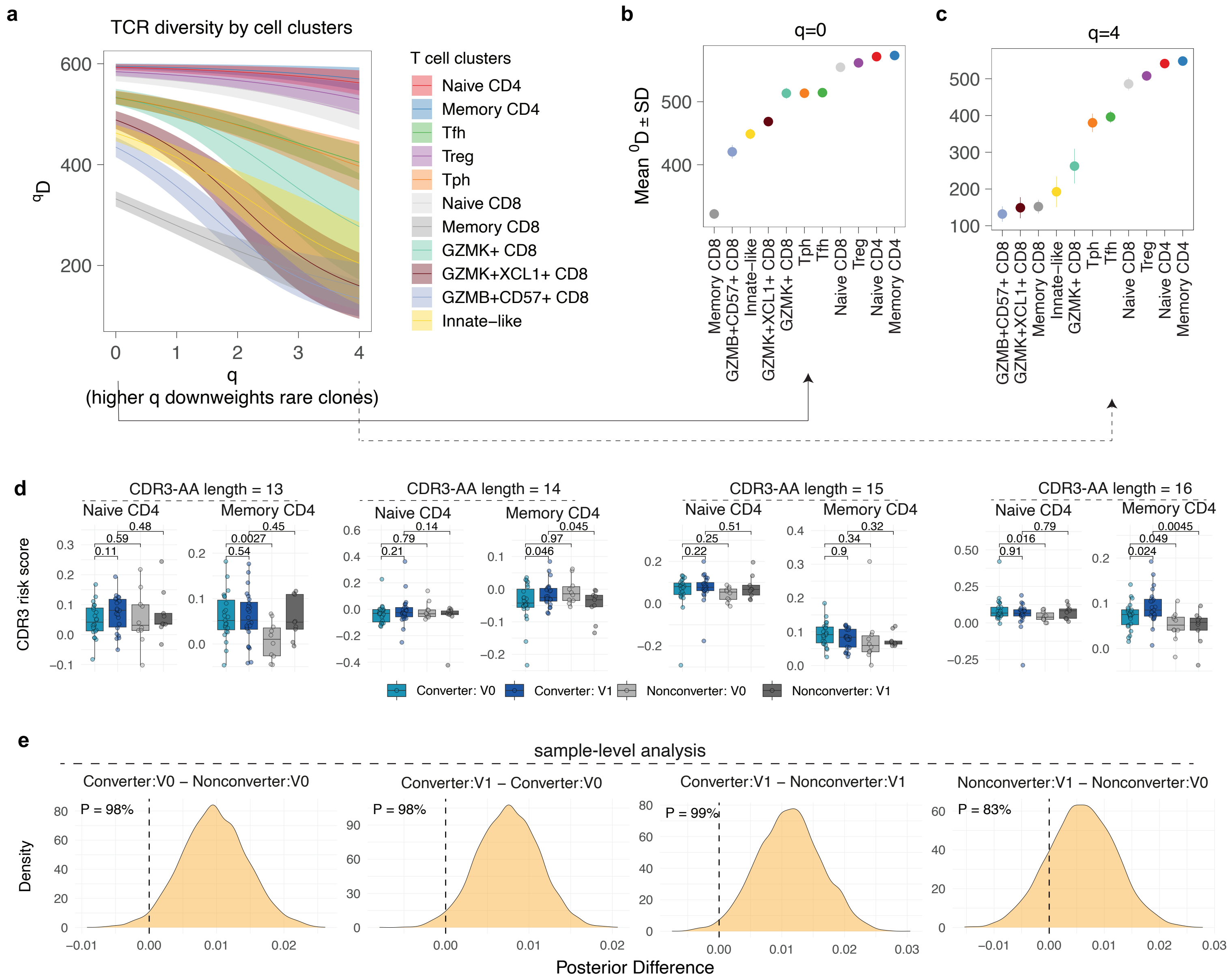
a
TCR diversity by cell clusters
T cell clusters
Naive CD4
Memory CD4
Tfh
Treg
Tph
Naive CD8
Memory CD8
GZMK+ CD8
GZMK+XCL1+ CD8
GZMB+CD57+ CD8
Innate−like
q
(higher q downweights rare clones)
b
q=0
Mean 0D ± SD
c
q=4
d
CDR3-AA length = 13
CDR3-AA length = 14
CDR3-AA length = 15
CDR3-AA length = 16
CDR3 risk score
Converter: V0
Converter: V1
Nonconverter: V0
Nonconverter: V1
e
sample-level analysis
Converter:V0 − Nonconverter:V0
Converter:V1 − Converter:V0
Converter:V1 − Nonconverter:V1
Nonconverter:V1 − Nonconverter:V0
P = 98%
P = 98%
P = 99%
P = 83%
Density
Posterior Difference

**Supplementary Figure 5: TCR repertoire analysis in CD4 and CD8 T cells. a**, TCR repertoire diversity as a smooth function (*D*) of a single parameter *q* by T cell clusters in the CITE-seq dataset. As the parameter *q* increases from 0 to + ∞ the diversity index (*D*) depends less on rare clones and more on common (abundant) ones. Large diversity index (*D*) is interpreted as high diversity in clonal populations. **b-c**, *D* zoomed at q=0 (**b**) and q=4 (**c**). **d**, Distribution of mean RA-CDR3 risk scores within naïve and memory $CD4^+$ T cell subsets, stratified by CDR3 AA length (13–16), for each sample. P-values represent results from one-sided t-tests. **e**, Sample-level posterior comparisons of RA-CDR3 scores between Converters and Nonconverters at baseline and follow-up accounting for variation by CDR3 length and participant. Each panel shows the posterior distribution of the difference between group means based on Bayesian modeling of single-cell RA-CDR3 scores accounting for variation by CDR3 length. Vertical dashed lines denote zero, and shaded densities represent the estimated distribution of group differences. The posterior probability P shown in each panel indicates the proportion of MCMC samples where the left group has higher scores than the right group.

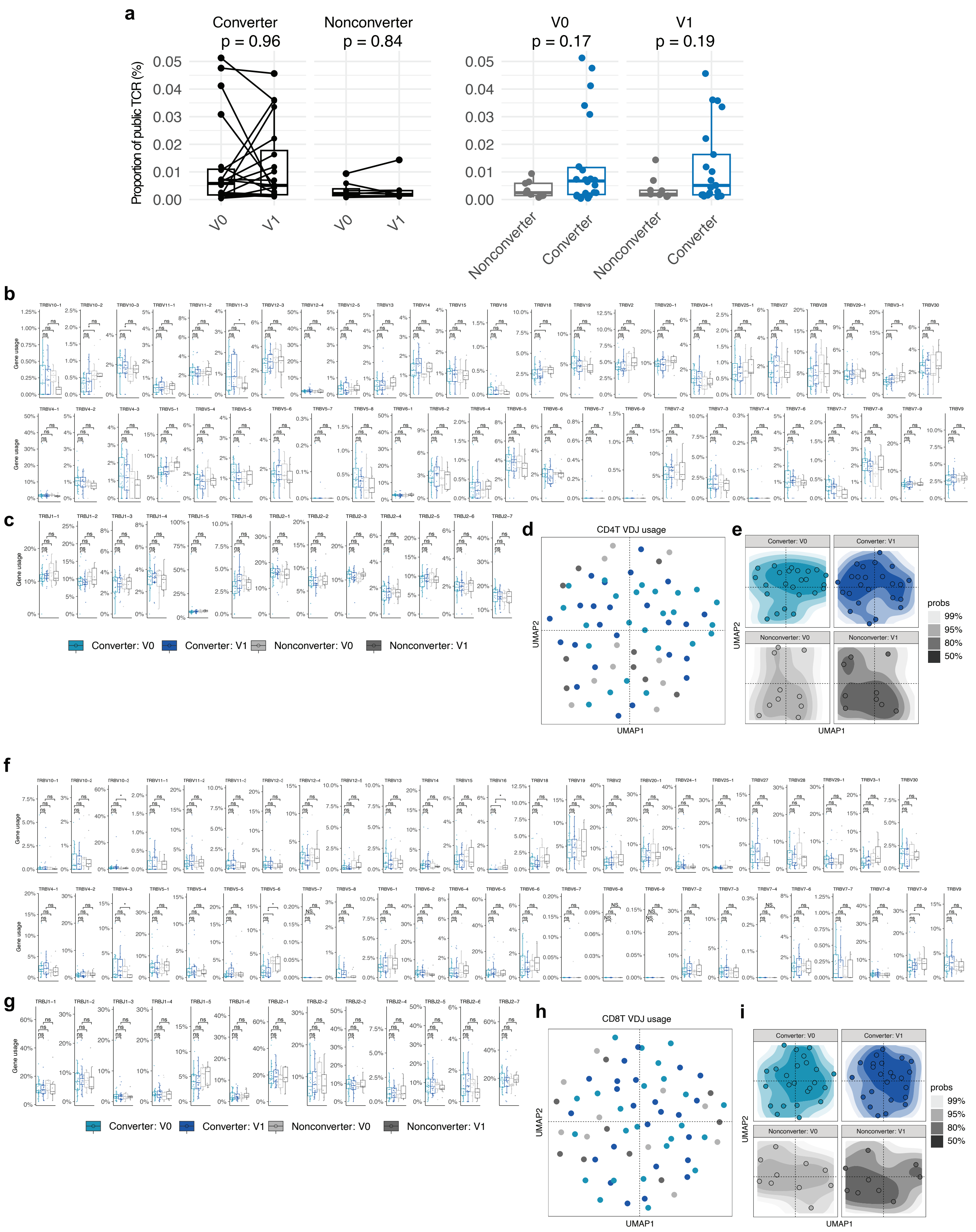
a
Converter
p = 0.96
Nonconverter
p = 0.84
V0
p = 0.17
V1
p = 0.19
Proportion of public TCR (%)
0.05
0.04
0.03
0.02
0.01
0.00
V0
V1
Nonconverter
Converter
b
Gene usage
c
Converter: V0
Converter: V1
Nonconverter: V0
Nonconverter: V1
d
CD4T VDJ usage
UMAP1
UMAP2
e
Converter: V0
Converter: V1
Nonconverter: V0
Nonconverter: V1
probs
99%
95%
80%
50%
f
g
h
CD8T VDJ usage
i

**Supplementary Figure 6: TCR usage analysis.** **a**, Paired line plots showing the percentage of public TCRs in Converters and Nonconverters at baseline (V0) and follow-up (V1). P-values from paired and unpaired Wilcoxon tests are indicated. **b-c**, Boxplots showing usage frequencies of individual TCR gene segments (TRBV and TRBJ respectively) across four groups: Converter V0, Converter V1, Nonconverter V0, and Nonconverter V1 for $CD4^+$ T cells. **d**, UMAP visualization of $CD4^+$ T cell samples colored by time point and converter status, based on VDJ usage. **e**, Kernel density estimation plots of UMAP in (**d**), showing the distribution probability of each group across UMAP space. **f–g**, Boxplots showing usage frequencies of individual TCR gene segments (TRBV and TRBJ respectively) across four groups: Converter V0, Converter V1, Nonconverter V0, and Nonconverter V1 for $CD8^+$ T cells. **h**, UMAP visualization of $CD8^+$ T cells based on VDJ usage across conditions. **i**, Kernel density estimation plots of UMAP in (**h**), showing the distribution probability for each group.

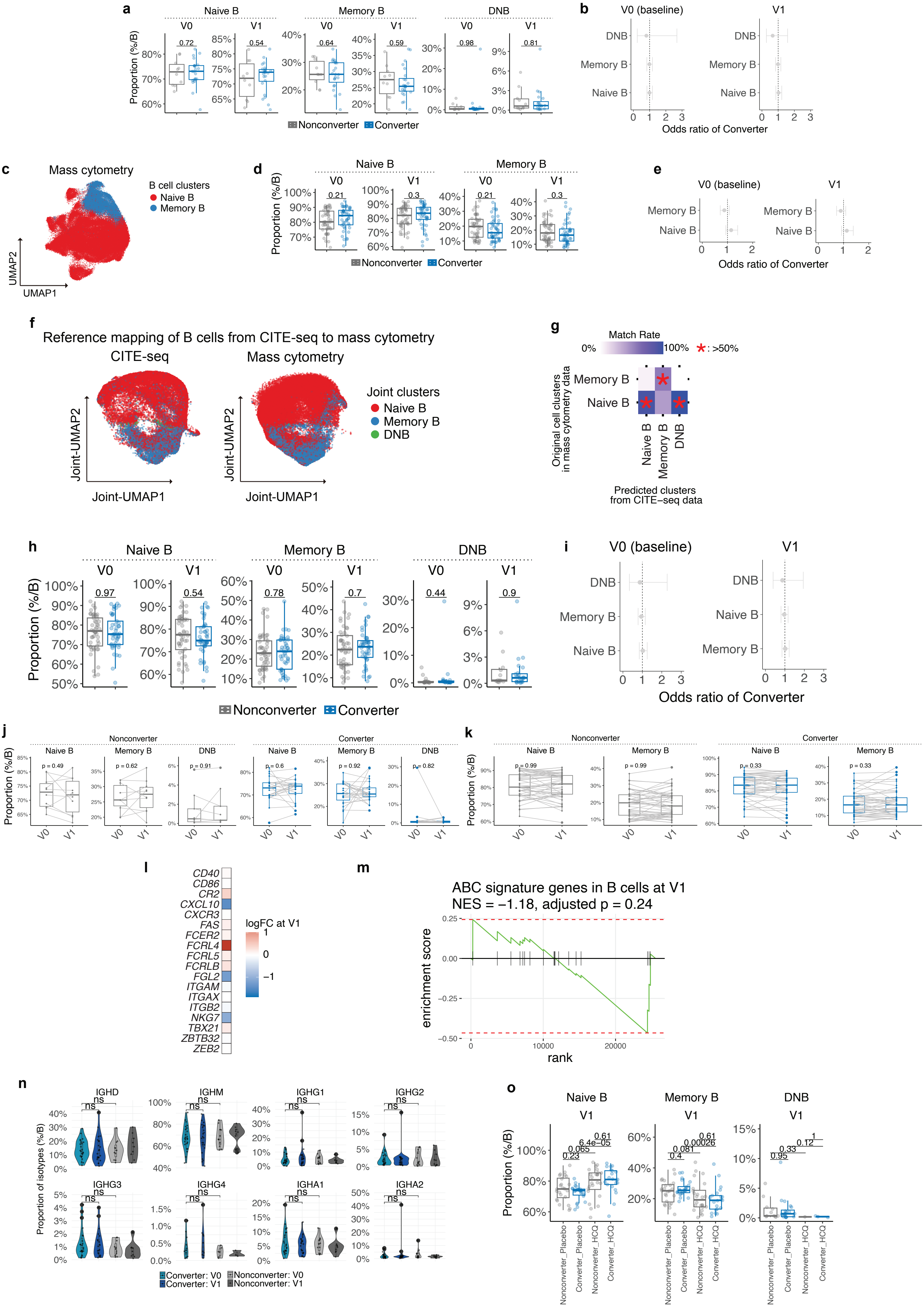
a
Naive B
Memory B
DNB
V0
V1
Proportion (%/B)
Nonconverter
Converter
b
V0 (baseline)
V1
DNB
Memory B
Naive B
Odds ratio of Converter
c
Mass cytometry
B cell clusters
Naive B
Memory B
UMAP1
UMAP2
d
e
f
Reference mapping of B cells from CITE-seq to mass cytometry
CITE-seq
Mass cytometry
Joint clusters
Joint-UMAP1
Joint-UMAP2
g
Match Rate
0%
100%
*: >50%
Original cell clusters in mass cytometry data
Predicted clusters from CITE−seq data
h
i
j
Nonconverter
Converter
k
l
logFC at V1
CD40
CD86
CR2
CXCL10
CXCR3
FAS
FCER2
FCRL4
FCRL5
FCRLB
FGL2
ITGAM
ITGAX
ITGB2
NKG7
TBX21
ZBTB32
ZEB2
m
ABC signature genes in B cells at V1
NES = −1.18, adjusted p = 0.24
enrichment score
rank
n
IGHD
IGHM
IGHG1
IGHG2
IGHG3
IGHG4
IGHA1
IGHA2
Proportion of isotypes (%/B)
Converter: V0
Converter: V1
Nonconverter: V0
Nonconverter: V1
o
Nonconverter_Placebo
Converter_Placebo
Nonconverter_HCQ
Converter_HCQ

**Supplementary Figure 7: B cell analysis using CITE-seq and mass cytometry data. a**, Proportion of each B cell cluster of CITE-seq data by time point and clinical outcome. **b**, GLMM analysis using CITE-seq B cells comparing RA Converters and Nonconverters at each visit point, at baseline (V0, left) and post-onset/follow-up (V1). The model accounted for age, sex, batch and individuals as covariates. The odds ratio and 95% confidence intervals (CIs) are shown by error bars. **c**, B cell clusters of mass cytometry dataset on UMAP. **d**, Proportion of each B cell cluster of mass cytometry data by time point and clinical outcome. **e**, GLMM analysis using mass cytometry B cells comparing RA Converters and Nonconverters at each visit point, at baseline (V0, left) and post-onset/follow-up (V1). The model accounted for age, sex, batch and individuals as covariates. The odds ratio and 95% confidence intervals (CIs) are shown by error bars. **f**, Joint UMAP showing integration of B cell data from CITE-seq (left) and mass cytometry (right), with reference mapping from CITE-seq clusters (Naive B, Memory B, DNB) to mass cytometry clusters. **g**, Heatmap showing match rates between predicted CITE-seq clusters and original mass cytometry-defined B cell clusters. Asterisks indicate matches with >50% overlap. **h**, Boxplots of subset proportions of joint B cell clusters in Converters and Nonconverters across V0 and V1. P-values were calculated using Wilcoxon rank-sum tests. **i**, GLMM analysis using joint B cell clusters comparing RA Converters and Nonconverters at each visit point, at baseline (V0, left) and post-onset/follow-up (V1). The odds ratio and 95% confidence intervals (CIs) are shown by error bars. **j-k**, Longitudinal comparison of B cell subsets frequencies in CITE-seq data (**j**) and mass cytometry data (**k**) between baseline (V0) and follow-up (V1) time points, stratified by RA conversion status. P-values from paired Wilcoxon signed-rank tests are indicated for each group. **l**, Heatmap of log fold changes at V1 between Converters and Nonconverters for representative ABC (age-associated B cell)-related genes. **m**, Gene set enrichment analysis (GSEA) of ABC signature genes at V1 in Converters versus Nonconverters. **n**, Violin plots showing proportions of isotypes (IGHM, IGHD, IGHGs, IGHAs) at V0 and V1 in Converters and Nonconverters. **o**, Cross-sectional comparison of B cell subset frequencies in joint space at follow-up (V1) between Converters and Nonconverters stratified by treatment arm (Placebo or HCQ). Boxplots display the proportions of B cell subsets within total B cells, respectively. Gray indicates Nonconverters, and blue indicates Converters. P-values were calculated using Wilcoxon rank-sum tests between groups.

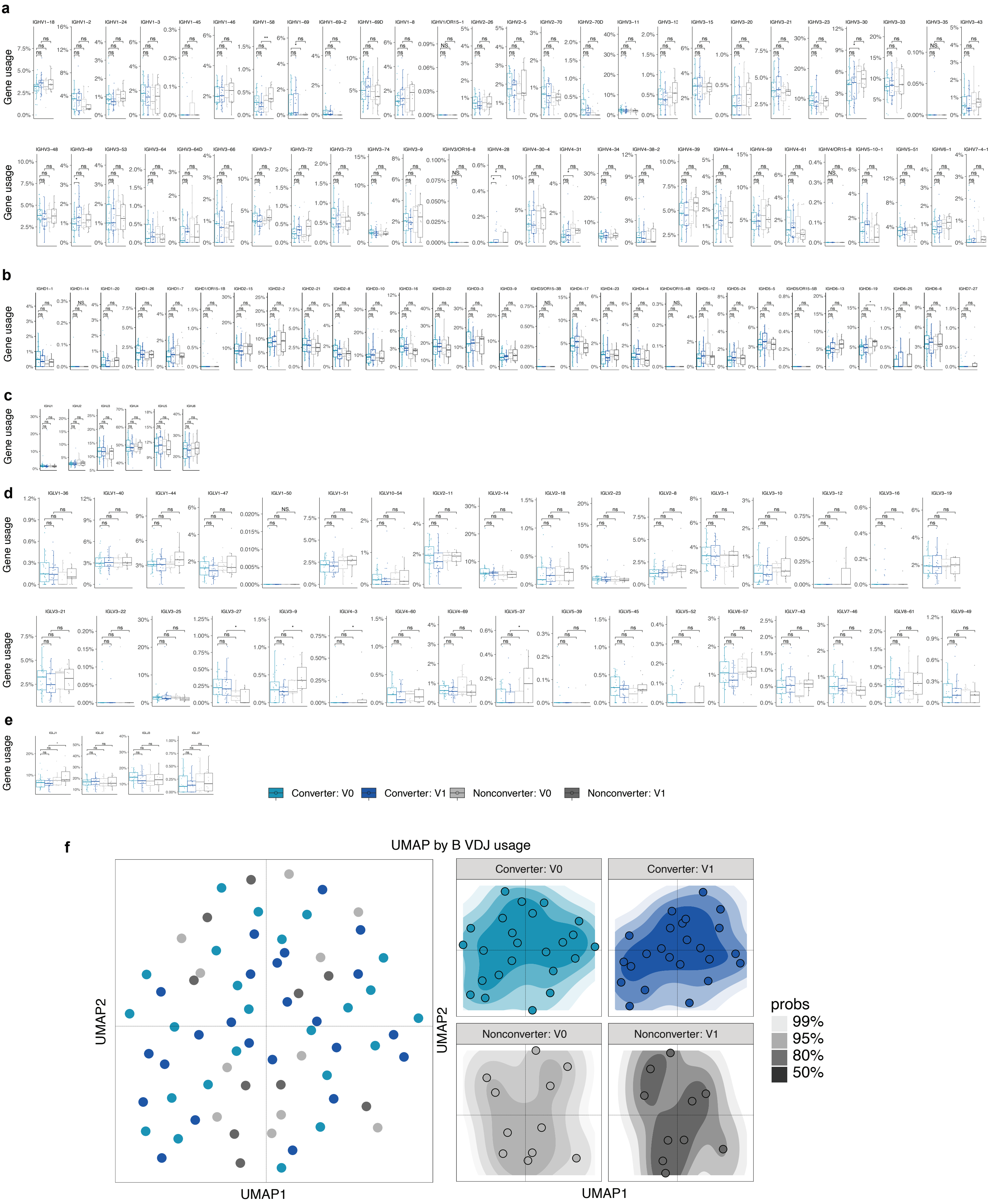
a
b
c
d
e
f
Gene usage
Converter: V0
Converter: V1
Nonconverter: V0
Nonconverter: V1
UMAP by B VDJ usage
UMAP1
UMAP2
probs
99%
95%
80%
50%

**Supplementary Figure 8: BCR usage analysis. a-e**, Boxplots of gene usage for IGH and IGL across groups; V gene in IGH (**a**), D gene in IGH (**b**), J gene in IGH (**c**), V gene in IGL (**d**), and J gene in IGL (**e**). **f**, UMAP of BCR VDJ usage across groups; density contour plots highlight distribution differences.

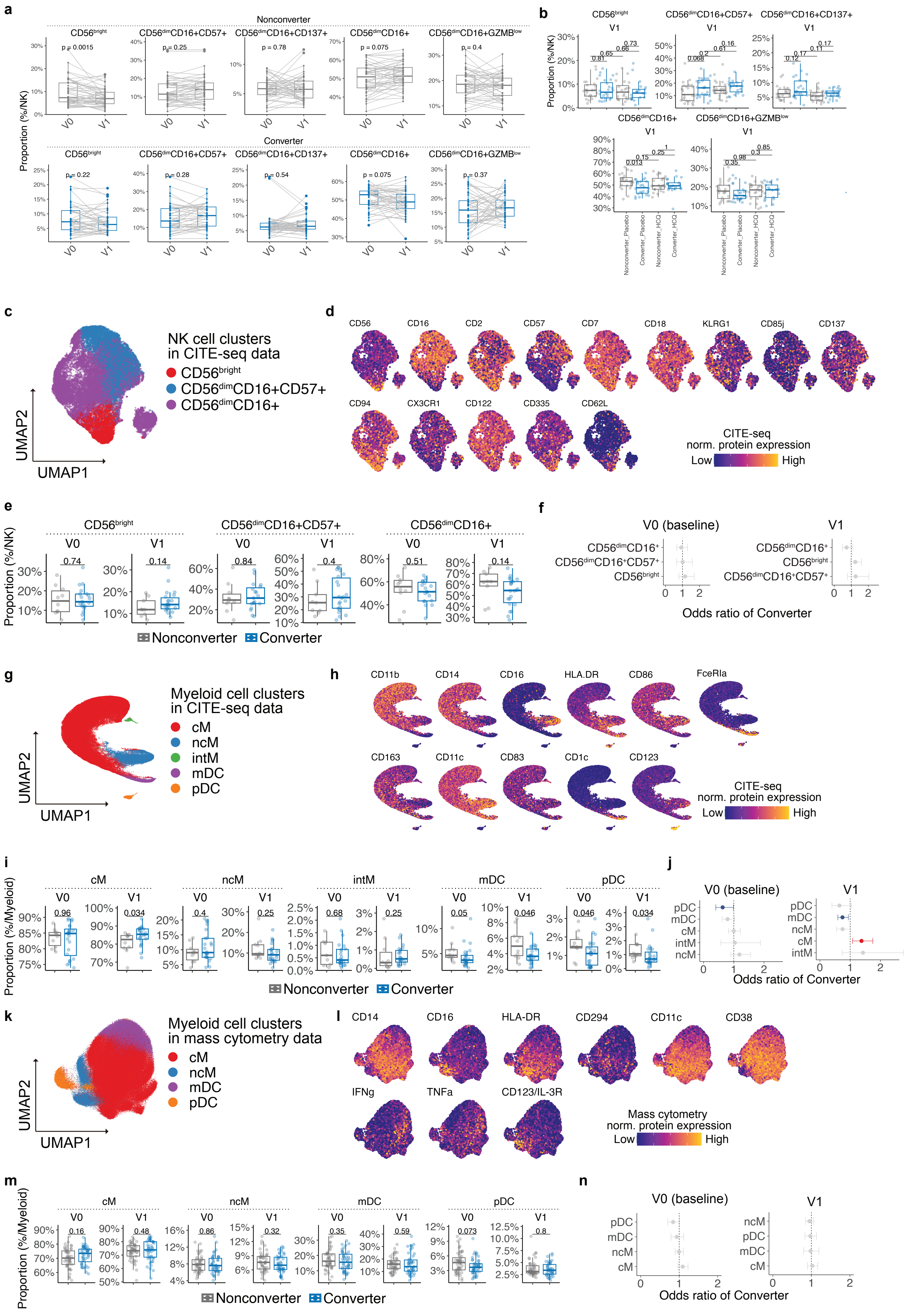

a
Nonconverter
Converter
CD56bright
CD56dimCD16+CD57+
CD56dimCD16+CD137+
CD56dimCD16+
CD56dimCD16+GZMBlow
Proportion (%/NK)
V0
V1
b
c
NK cell clusters
in CITE-seq data
CD56bright
CD56dimCD16+CD57+
CD56dimCD16+
UMAP1
UMAP2
d
CD56
CD16
CD2
CD57
CD7
CD18
KLRG1
CD85j
CD137
CD94
CX3CR1
CD122
CD335
CD62L
CITE-seq
norm. protein expression
Low
High
e
Nonconverter
Converter
f
V0 (baseline)
V1
Odds ratio of Converter
g
Myeloid cell clusters
in CITE-seq data
cM
ncM
intM
mDC
pDC
h
CD11b
CD14
CD16
HLA.DR
CD86
FceRIa
CD163
CD11c
CD83
CD1c
CD123
i
Proportion (%/Myeloid)
j
k
Myeloid cell clusters
in mass cytometry data
l
CD14
CD16
HLA-DR
CD294
CD11c
CD38
IFNg
TNFa
CD123/IL-3R
Mass cytometry
norm. protein expression
m
n

**Supplementary Figure 9: Integrated analysis of NK and myeloid cells from CITE-seq and mass cytometry data. a**, Paired comparison of NK cell subset frequencies in mass cytometry data at baseline (V0) and post-onset/follow-up (V1) within Nonconverters (top) and Converters (bottom). P-values were calculated by Wilcoxon signed-rank tests. **b**, Frequencies of NK cell subsets at V1, stratified by clinical outcome and treatment group (placebo or HCQ). P-values were calculated by Wilcoxon rank-sum tests. **c**, UMAP visualization of NK cell clusters identified from CITE-seq data, annotated as $CD56^{bright}$, $CD56^{dim}CD16^{+}CD57^{+}$, and $CD56^{dim}CD16^{+}$ subsets. **d**, Expression levels of selected NK-associated surface proteins projected onto the CITE-seq UMAP. **e**, Comparison of NK subset frequencies between Converters and Nonconverters at baseline (V0) and follow-up (V1). P-values from Wilcoxon tests are shown above each boxplot. **f**, GLMM analysis using CITE-seq NK cells comparing RA Converters and Nonconverters at each visit point, at baseline (V0, left) and post-onset/follow-up (V1). The odds ratio and 95% confidence intervals (CIs) are shown by error bars. **g**, UMAP visualization of myeloid cell clusters from CITE-seq data, including classical monocytes (cM), non-classical monocytes (ncM), intermediate monocytes (intM), myeloid dendritic cells (mDC), and plasmacytoid dendritic cells (pDC). **h**, Protein expression levels of selected myeloid markers across CITE-seq-defined myeloid populations. **i**, Boxplots comparing myeloid subset proportions between Converters and Nonconverters at V0 and V1. Significant differences are highlighted with asterisks. **j**, GLMM analysis using CITE-seq myeloid cells comparing RA Converters and Nonconverters at each visit point, at baseline (V0, left) and post-onset/follow-up (V1). The odds ratio and 95% confidence intervals (CIs) are shown by error bars. **k**, UMAP showing myeloid cell clusters in mass cytometry data, including cM, ncM, mDC+mixed, and pDC subsets. **l**, Protein expression of canonical myeloid markers in mass cytometry data projected onto UMAP. **m**, Proportions of each myeloid cell subset in Converters and Nonconverters at V0 and V1 in mass cytometry data. **n**, GLMM analysis using mass cytometry myeloid cells comparing RA Converters and Nonconverters at each visit point, at baseline (V0, left) and post-onset/follow-up (V1). The odds ratio and 95% confidence intervals (CIs) are shown by error bars.

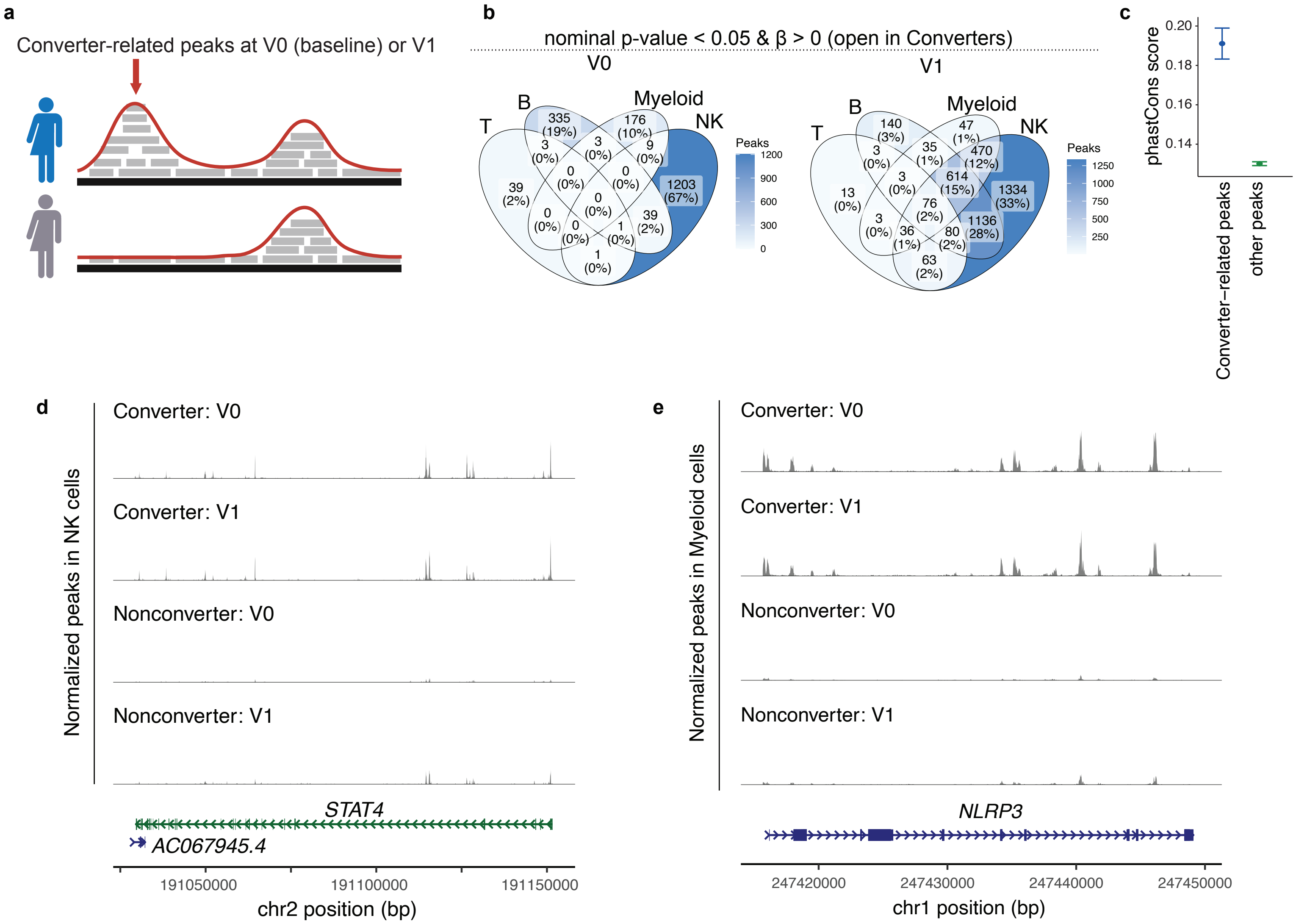
a
Converter-related peaks at V0 (baseline) or V1
b
nominal p-value < 0.05 & β > 0 (open in Converters)
V0
V1
T
B
Myeloid
NK
Peaks
335 (19%)
176 (10%)
1203 (67%)
39 (2%)
140 (3%)
47 (1%)
470 (12%)
614 (15%)
1334 (33%)
1136 (28%)
c
phastCons score
Converter−related peaks
other peaks
d
Normalized peaks in NK cells
Converter: V0
Converter: V1
Nonconverter: V0
Nonconverter: V1
STAT4
AC067945.4
191050000
191100000
191150000
chr2 position (bp)
e
Normalized peaks in Myeloid cells
Converter: V0
Converter: V1
Nonconverter: V0
Nonconverter: V1
NLRP3
247420000
247430000
247440000
247450000
chr1 position (bp)

**Supplementary Figure 10: Chromatin accessibility changes in immune cells of Converters.** **a**, Schematic illustrating the detection of chromatin peaks enriched in Converters compared to Nonconverters, Converter-related peak, at baseline (V0) or post-onset/follow-up (V1). **b**, Venn diagrams showing the overlap of Converter-enriched peaks among immune cell types at V0 and V1. Numbers represent peak counts and percentages of total Converter-related peaks. **c**, Mean ± 1.96 × Standard error of the phastCons scores. **d-e**, Representative tracks showing accessibility signals in Converters and Nonconverters across time points at selected loci: (**d**) Locus around *STAT4* (NK cells) and (**e**) Locus around *NLRP3* (Myeloid cells).

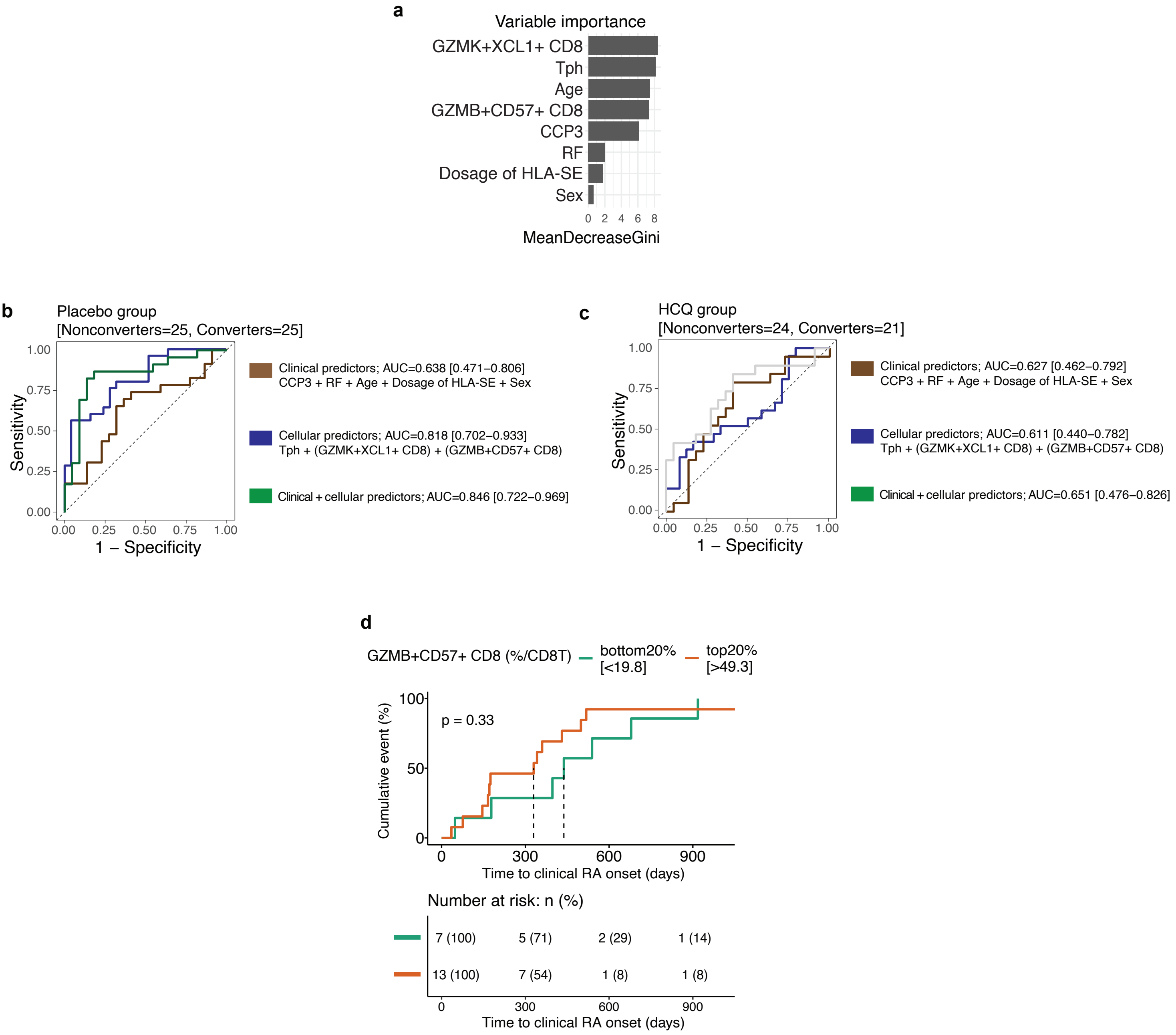
a
Variable importance
GZMK+XCL1+ CD8
Tph
Age
GZMB+CD57+ CD8
CCP3
RF
Dosage of HLA-SE
Sex
0 2 4 6 8
MeanDecreaseGini
b
Placebo group
[Nonconverters=25, Converters=25]
Sensitivity
1.00
0.75
0.50
0.25
0.00
0.00 0.25 0.50 0.75 1.00
1 – Specificity
Clinical predictors; AUC=0.638 [0.471–0.806]
CCP3 + RF + Age + Dosage of HLA-SE + Sex
Cellular predictors; AUC=0.818 [0.702–0.933]
Tph + (GZMK+XCL1+ CD8) + (GZMB+CD57+ CD8)
Clinical + cellular predictors; AUC=0.846 [0.722–0.969]
c
HCQ group
[Nonconverters=24, Converters=21]
Sensitivity
1.00
0.75
0.50
0.25
0.00
0.00 0.25 0.50 0.75 1.00
1 – Specificity
Clinical predictors; AUC=0.627 [0.462–0.792]
CCP3 + RF + Age + Dosage of HLA-SE + Sex
Cellular predictors; AUC=0.611 [0.440–0.782]
Tph + (GZMK+XCL1+ CD8) + (GZMB+CD57+ CD8)
Clinical + cellular predictors; AUC=0.651 [0.476–0.826]
d
GZMB+CD57+ CD8 (%/CD8T)
bottom20%
[<19.8]
top20%
[>49.3]
p = 0.33
Cumulative event (%)
100
50
0
0 300 600 900
Time to clinical RA onset (days)
Number at risk: n (%)
7 (100) 5 (71) 2 (29) 1 (14)
13 (100) 7 (54) 1 (8) 1 (8)
0 300 600 900
Time to clinical RA onset (days)

**Supplementary Figure 11: Evaluation of predictors for time to RA onset. a,** Variable importance plot from a random forest classifier evaluating predictors of progression to clinical RA. The model incorporated both clinical and cellular features. **b-c**, Receiver operating characteristic (ROC) curves of the placebo group (**b**) and HCQ group (**c**) comparing the performance of models based on (1) Clinical predictors (brown), (2) Cellular predictors (blue), and (3) Combined clinical + cellular predictors (green). **d,** Kaplan–Meier survival analysis showing time to clinical RA onset stratified by the proportion of *GZMB*$^{+}$*CD57*$^{+}$ CD8+ T cells at baseline (V0).
